# Neighborhood Influences on the Geography of Type 2 Diabetes in Malaysia: A Geospatial Modelling Study

**DOI:** 10.1101/2024.10.26.24316183

**Authors:** Kurubaran Ganasegeran, Mohd Rizal Abdul Manaf, Nazarudin Safian, Lance A. Waller, Feisul Idzwan Mustapha, Khairul Nizam Abdul Maulud, Muhammad Faid Mohd Rizal

**Affiliations:** Department of Public Health Medicine, Faculty of Medicine, Universiti Kebangsaan Malaysia, 56000 Kuala Lumpur, Malaysia; Occupational Safety and Health Unit, Seberang Jaya Hospital, Ministry of Health Malaysia, 13700 Seberang Perai, Penang, Malaysia; Public Health Unit, Seberang Jaya Hospital, Ministry of Health Malaysia, 13700 Seberang Perai, Penang, Malaysia; Department of Biostatistics and Bioinformatics, Rollins School of Public Health, Emory University, Atlanta, GA 30322, USA; USA; Public Health Division, Perak State Health Department, Ministry of Health Malaysia, 30000 Perak, Malaysia; Earth Observation Centre (EOC), Institute of Climate Change, Universiti Kebangsaan Malaysia, 43600 Selangor Darul Ehsan, Malaysia; Department of Civil Engineering, Faculty of Engineering & Built Environment, Universiti Kebangsaan Malaysia, 43600 Selangor Darul Ehsan, Malaysia

**Keywords:** type 2 diabetes, neighborhood, geospatial, geography, population, public health policy

## Abstract

Type 2 diabetes (T2D) often exhibits long-standing disparities across populations. Spatial regression models can identify areas of epidemiological conformity and transitions between local neighborhoods to inform timely, localized public health interventions. We identified areal-level distributions of T2D rates across Malaysia and synthesized prediction models to estimate local effects and interactions of different neighborhood covariates affecting local T2D burden. We obtained aggregated counts of national level T2D cases data by administrative-districts between 2016-2020 and computed district-wise crude rates to correlate with district-level neighborhood demographic, socio-economic, safety, fitness, access to built-environments, and urban growth indicators from various national sources and census data. We applied simultaneous spatial autoregressive (SAR) models coupled with two-way interaction analyses to account for spatial autocorrelation and estimate risk factors for district-level T2D rates in Malaysia. The variation in spatial lag estimates of T2D rates by districts was influenced by the proportion of households living below 50% of the median income (β = 0.009, *p* = 0.002) and national poverty line (β = - 0.012, *p* = 0.001), income inequalities (β = - 2.005, *p* = 0.004), CCTV coverage per 1000 population (β = 0.070, *p* = 0.023), average property crime index per 1000 population (β = 0.014, *p* = 0.033), access to bowling centers (β = - 0.003, *p* = 0.019), and parks (β = 0.007, *p* = 0.001). Areal-level district-wise crude T2D rate estimates were influenced by neighborhood socio-economic vulnerabilities, neighborhood safety, and neighborhood access to fitness facilities, after accounting for residual spatial correlation via SAR models.

## Introduction

Neighborhoods are places with social and cultural meaning to local residents, defining where they typically reside, work, play, or have a sense of belonging.^1,2^ Neighborhoods are built based on people’s living needs, expectations, and circumstances, alongside with urbanization processes offering basic physical and landscape features (e.g., parks, sidewalks, or bicycle lanes), resulting from local township planning, maintenance, or restructuring.^2^ The ultimate target of such interventions is to achieve sustainable neighborhood outcomes for livability, equity, and viability.^3^ More generally, neighborhoods reflect geographical entities of smaller confined residential areas or human settlements, nested within larger units like cities, states, or regions.^2^

With respect to neighborhood operations and type 2 diabetes (T2D), local risks often are based on local upstream social, political, or commercial drivers influencing local community health throughout the life course^4^ – e.g., as measured by the rise of type 2 diabetes (T2D) or obesity burdens with evidence of longstanding disparities between sociodemographic sub-populations.^5,6^ Theoretical frameworks postulate contemporary neighborhoods being susceptible to T2D risk owing to limited access to healthy food, and the influence of proximity or density metrics to fitness facilities or green spaces – and when coupled with poverty, local crimes, social delinquency, local colonialism, or globalization processes, these neighborhood environments cause potential psychosocial stress to local residents, prompting a decline of lifestyle behaviors, and resulting greater susceptibility to T2D risks or other chronic conditions.^6,7^

Socio-economic vulnerabilities such as poverty, income inequalities, and unemployment rates have been positively linked to various lifestyle and chronic health conditions.^7-11^ These established linkages were mostly interpreted at the individual level across the literature without considering the specific geographical scales capable of providing a snapshot to local population risks for public health interventions. Areal-level economic markers such as the Gini coefficient offer measures of income inequalities (i.e., the extent of household income distributed unevenly) captured by national population censuses that are useful for understanding local landscapes of human behavior and living circumstances across communities - where the rich are often clustered in areas of decent living, while the poor typically struggle to survive within the social and commercial environmental hazards – often predisposing them to greater health needs within chaotic neighborhoods.^3,12,13^

The rural-urban matrix coupled with each neighborhood’s demography (i.e., ethnic/racial disparities or minorities; women; middle-to-older aged people) and neighborhood’s socio-economic vulnerabilities often reveal significant correlations with high neighborhood-level obesity and T2D rates.^8,14^ These vulnerable groups are often segregated or pressured in deprived neighborhoods, accelerating their neighborhood safety concerns due to higher incidence of social delinquency, local crime, or drug addiction – a phenomenal tendency towards increasing psychological stress and T2D risk.^15,16^

Local safety concerns may decrease physical activity - a cornerstone for T2D risk management - among residents dwelling within deprived neighborhoods, likely due to reduced walking, cycling, or access to nearby residential built-environments for exercise or recreational activities (e.g., neighborhood parks, sports, or fitness facilities).^14,16^ In addition, physical inactivity often is connected to people’s demographic, socio-economic characteristics, and health outcomes, where ethnic/racial minorities or women from lower income groups in urban areas often exhibit greater risks for poorer health, especially in rural areas.^17,18^ Some characteristics of human settlements, such the percentage living in rural areas are known to lower rates of physical inactivity as compared to their urban counterparts,^19^ thus accelerating risk for chronic diseases among local residents.^16,20^

Consistent with the regional atlas estimates illustrating that Malaysia topped T2D prevalence among countries in the Western Pacific Region as of 2021,^21^ the National Health and Morbidity Survey (NHMS) 2019 Report claimed that approximately 1 in 5 adults in Malaysia was afflicted with T2D, totalling to approximately 3.9 million people above the age of 18 years.^22^ Countrywide point prevalence of T2D increased from 11.2% in 2011 to 18.3% in 2019.^22^

The burden of T2D escalated alongside demographic, societal, and cultural changes within Malaysian communities. Common individual-level risk factors for T2D generally include physical inactivity, poor dietary behaviours, and demographic profiles such as age, ethnicity, socio-economic status, or type of occupations.^23-26^ But these attributes often associate with neighborhood communities,^27^ where local social determinants of health can catalyse poor health amongst different residential areas.^28^ Local variations in local health are greatly influenced by social stratifications, causing disparities in T2D prevalence rates and associated risk factors, all of which can be influenced by *“place.”*

Here, we explore the distribution of T2D crude-rates by administrative districts in Malaysia, and subsequently determine the influence of local demography, socio-economic vulnerabilities, safety, urban growth indicators, neighborhood fitness, and neighborhood access to built-environments on the geography of T2D. As neighborhoods are comprised of multiple structural and social determinants shaping the landscape of human living conditions across geography, we further examine potential interaction effects between selected neighborhood covariates and the spatial variation of T2D rates. Our study assessed a wide range of neighborhood indicators complementing the attributes outlined within the Sustainable Development Goals (SDG) framework for healthy and dignified living.

## Methods

### Study Design, Setting, and Population

For this ecological study, we assembled and linked district-level population data (n = 144), countrywide, for Malaysia, using multiple data sources from the National Diabetes Register (NDR),^29^ administrative shapefiles (level 1 for states; level 2 for administrative districts),^30^ and population wellbeing surveys and social indicators integrated within the Malaysian Population Census.^31^ Data on 271,553 active T2D adults aged ≥ 20 years from 2016 to 2020 by administrative-districts were retrieved from the National Diabetes Register (NDR) of Malaysia.^29^ Active T2D cases diagnosed based on local clinical practice guidelines^32^ with at least one follow-up visit to the registered primary health clinic in a year were captured.^33^ In line with universal healthcare coverage, primary health services in Malaysia serve a catchment area within five kilometers of community neighborhoods, which are typically nested within local administrative-districts, as principally captured within the NDR.

### Measures

Our main outcome measure was district-wise crude rates of T2D computed from aggregated cases reported to the NDR. We estimated associations between local T2D rates and covariates consisting of neighborhood demography (e.g., proportions of ethnic minorities), neighborhood socio-economic vulnerability (e.g., income inequalities), urban growth, neighborhood safety (e.g, property crime indices), and proximity to built-environment opportunities for fitness activities (e.g., access to parks or sports facilities).

### Metrics and Data Sources

Covariates retrieved from different data sources are described as follows:

i. *Diabetes rates* **-** We computed T2D crude prevalence rates per 100,000 population based on the proportion of cumulative cases from the years 2016-2020 to the total adult population aged ≥ 20 years for each district in Malaysia [total population of adults aged ≥ 20 years approximated to 19,697,300 people countrywide, consisting of 144 districts^31^, a common measure allowing comparisons across different regions or areas irrespective of population size and interpretating coefficients synthesized from regression models^34^.
ii. *Neighborhood’s demography* **-** In line with global trends^35-39^, the Malaysian National Health and Morbidity Survey (NHMS 2019) reported that T2D was highly prevalent in women, ethnic minority (Indians) followed by ethnic Bumiputera, and advancing age^22^. At the district-level, we operationalized neighborhood’s demography of adults aged ≥ 20 years as continuous variables based on the proportion of women and men, minority ethnic composition (proportion of Indians), proportion of Bumiputera (i.e., Bumiputera Malay, Bumiputera Others), proportion of Chinese, and the proportion of adults aged 20-34 years, 35-49 years, 50-64 years, and ≥65 years, retrieved from the Malaysian population census^31^.
iii. *Neighborhood’s socio-economic vulnerability* **-** Neighborhood’s socio-economic vulnerability was measured through three domain indicators of poverty, income inequalities, and unemployment. Poverty was measured based on the United Nations Sustainable Development Goals (Goal 1: No Poverty) 2019 indicator that reports the proportion of households living below the national poverty line (poverty line income for Malaysia in 2019 was MYR 2208 per month; conversion based on 2019 average exchange rate was USD 533.10) by administrative districts from the Malaysian population census^31^. Additionally, poverty was also measured based on the proportion of people living below fifty percent of median income by administrative districts from the Malaysian population census^31^. This metric is defined as the percentage of people in the population by administrative districts who live in households whose per capita income or consumption is below half of the median income (median household income for Malaysia in 2019 was MYR 5873 per month; conversion based on 2019 average exchange rate was USD 1417.98; below 50% of median income equals to MYR 2937.50 or less per month; conversion based on 2019 average exchange rate was USD 709.54) or consumption per capita. The median was measured based on the 2017 Purchasing Power Parity (PPP) using the Poverty and Inequality Platform from the World Bank^40^. While the poverty line defines a measure defining a state of adequate requirement of income by an individual to fulfill the basic needs of livelihood (as determined by national policy), the current study added the “below 50% of median income” as second, alternative measure of local poverty based on local concentrations of individuals in the lower quarter of the national income distribution. This covariate is a relative measure of poverty which could change considerably overtime based on country’s economic performance, as poverty thresholds could rise or decline rapidly during periods of economic growth or downturn, affecting growth or development of neighborhoods social structure within urbanization processes. The two measures (included separately in our regression models) represent different aspects of local poverty and comparisons of results can provide more nuanced insight into the types of associations between T2D and income^41^. Local income *inequality* was conceptualized based on the United Nations Sustainable Development Goals (Goal 10: Reducing Inequalities) 2019 indicator as measured by local Gini coefficients in districts from the Malaysian population census^31^. A local Gini coefficient of 0 indicates perfect income equality within the subregion and 1 (or 100) reflects maximal income inequality^40^. Unemployment rates by administrative districts was measured with reference to the United Nations Sustainable Development Goals (Goal 8: Decent Work and Economic Growth) indicator, retrieved from the Malaysian population census^31^.
iv. *Urban growth indicator -* Urbanization processes would forcibly displace persons through internal migration, resulting in wider urban-rural gap of human settlements that influences township planning or restructuring. We measured urbanization processes that affect neighborhoods based on urban growth indicator. Urban growth rate was measured as the difference of urban population shift between the years of 2010 and 2020 in Malaysia retrieved from the Malaysian population census^31^.
v. *Neighborhood’s safety -* Neighborhood safety was conceptualized based on the United Nations Sustainable Development Goals (Goal 16: Peace, Justice, and Strong Institutions) indicator; defined as the proportion of population that feel safe walking alone around the neighborhood they live, while motivating local communities to access nearby public area facilities for engagement in fitness, physical, or sports activities^31^. Neighborhood safety was measured based on a proxy variable sourced from the 2021 Ministry of Housing and Local Government Authority social population statistics that computes the number of closed-circuit television (CCTV) cameras installed in a local authority area by administrative districts in Malaysia^31^. We further computed neighborhood safety as the number of CCTV coverage per 1000 population and acknowledged the effectiveness of the coverage for enhancing safety in deprived neighborhoods^42^. We computed average property crime index per 1000 population between the years 2018 and 2020 by administrative districts, sourced from the Royal Malaysia Police data, available through Malaysian population census^31^. Property crime index was defined according to the Standing Order of the Inspector General of Police (PTKPN) D203 that include frequently reported cases with sufficient significance to be considered as an important indicator; these include crimes related to house break-ins and thefts, vehicle thefts (e.g., van, lorry, motorcar, motorcycle), and other thefts (e.g., pick pocket, bicycle theft, public property theft)^31^. We calculated average drug addicts per 1000 population between the years 2018 and 2020 by administrative districts, sourced from the National Anti-Drugs Agency Malaysia data, available through the Malaysian population census^31^. Drug addicts were contextualized as per case data (i.e., those who have one or more offences in the current year). It refers to the psychoactive chemicals used (excluding alcohol, tobacco, and inhalants) not for medical purposes where their usage is prohibited, and that these substances lead to increased physical and psychological dependence and tolerance causing adverse effects on health, self, family, and society^31^. These crimes or addiction activities are vulnerable to cause societal chaos, vandalism, public area property dysfunction, or safety issues within disordered neighborhoods^43,44^.
vi. *Neighborhood fitness indicator and built-environment proximity -* Neighborhood fitness indicator by administrative-districts was assessed with a continuous variable based on the proportion of adult population aged ≥ 20 years not engaged in fitness, exercise, or sports activities, retrieved from the Population Wellbeing Fitness Survey^31^. The coverage of built-environments within neighborhoods were assessed through the availability of public facilities for communities’ fitness, recreational, exercise, or sports activities. This indicator was measured as the percentage of neighborhood areal access to recreational parks, jogging tracks, cycling tracks, mini stadiums, football fields, gymnasium, and bowling centers within five kilometers of proximity from resident’s home, which serves as a proxy for fitness, exercise, or physical activities available to local neighborhood residents. The rationale for the inclusion of different covariates on the types of sports, exercise, fitness, or recreational facilities in our model was based on the compositional structure of neighborhoods demography, as human settlements are ultimately composed of communities with different age groups. As a result, different sports or fitness activities attract residents of different age motivated to engage in different activity levels within their local neighborhoods. For example, members of older age groups often are likely to be engaged in light intensity physical activity such as walking, thus would seek access to recreational parks; whereas younger to middle aged groups are likely to be engaged in moderate to vigorous intensity physical activity such as running or jogging, bowling, running on a treadmill, soccer, or cycling, thus would likely access jogging tracks, cycling tracks, mini stadiums, football fields, gymnasium, or bowling centers^45^. Neighborhood sports and recreational facilities infrastructure development falls primarily under the jurisdiction of local municipalities. They are structured based on the fundamentals of “sports place theory,” a subset of the “central place theory” that correlates the provision of sports or recreational facilities with the level or urbanization processes within neighborhoods (i.e. greater urbanization have higher land use for the population, thus catalyze greater development of sports or fitness infrastructures for healthy urbanism)^46-48^. The national level cut-off points metrics of accessibility to sports and recreational facilities of at least five kilometers from residence home retrieved for this study represents a standard measure used in township development by local municipalities, a measure that also accounts for neighborhood demographics, urbanization processes, and neighborhood socioeconomic statuses for different types of sports infrastructure, as reported in previous work^49,50^. Our data for access to sports, fitness, or recreational facilities were sourced from the Population Wellbeing Fitness Survey^31^.

### Data Analysis

We conducted a comprehensive analysis involving the associations between T2D rates and neighborhood indicators across administrative districts. Following a summary of descriptive statistics for all variables used in our analysis, we executed exploratory spatial data analyses (ESDA), non-spatial correlations, spatial autocorrelations, and spatial regression modelling to evaluate the relationships between neighborhood-level attributes and T2D rates. We evaluated the distribution of crude rates via Q-Q plots if they were normally distributed, and if not, they were log-transformed to meet the assumption of a Gaussian distribution for spatial model fitting. As crude rates were subjected to variance instability that leads to spurious outliers, we performed rate smoothing via Empirical Bayes approach via GeoDA version 1.18 software (Center for Spatial Data Science University of Chicago, IL, USA) to visualize T2D distribution via a smoothed rate map.

### Cartographic Visualizations

We performed Geographic Information System (GIS) linkage through overlay of state boundaries and administrative districts through shapefiles, and spatially joined attribute data of various data sources from multiple agencies. ESDA involved cartographic development of a quantile map using Quantum GIS (QGIS), version 3.22 Bialowieza. This enabled geo-visualization of the distribution of T2D by administrative districts in Malaysia. Bivariate choropleth maps were built (via plugins ‘Bivariate Legend’) in QGIS using *n^2^* classes (three categories for each covariate yielding a total of nine categories) to map the combinations of statistically significant covariates associated with diabetes rates across geographies from the spatial regression models. The results were expressed as tertile groups, meaning 33% of the districts were classified in each of the “low,” “medium,” and “high” categories.

### Non-spatial Correlations and Spatial Autocorrelations

We conducted non-spatial analysis using Pearson correlations (*r*) between logged T2D rates and neighborhood indicators measured at the same area (within districts) using SPSS software version 23.0 (IBM). We then computed Global Moran’s *I* indexes to inspect the spatial autocorrelation of the outcome and of the covariates being tested, using first-order Queen’s contiguity spatial weights matrix that define neighbors as administrative-districts sharing a common border or corner using GeoDA version 1.18 software (Center for Spatial Data Science University of Chicago, IL, USA).

We determined Queen’s contiguity as the best spatial weight matrix for our work after running a series of twenty matrices for local clustering of T2D rates (i.e., Queen contiguity first and second order; Rook contiguity first and second order; distance matrix 150, 200, 250, 300 kilometers; inverse distance 150, 200, 250, 300 kilometers; k-nearest neighbor where k=2, 3, 4, 5; and k-nearest neighbor inverse where k=2, 3, 4, 5).

The Moran’s *I* value ranges approximately between -1 (negative spatial autocorrelation, showing neighborhood areas have dissimilar pattern) and 1 (positive spatial autocorrelation, suggesting neighborhood areas clustering with similar high or low values). A Moran’s *I* value near zero suggests complete spatial randomness. Moran’s *I* is expressed as follows^51,52^:

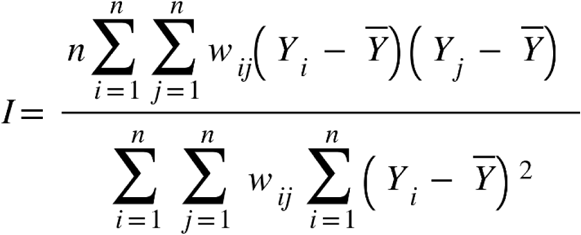

where *Y_i_* denotes outcome of interest (i.e., T2D rates) in area *i,* 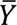 is the mean of covariate of interest, and *w_ij_* is an *n × n* spatial weight matrix that measures the “closeness” between area *i* and its neighbor *j*.

The adjacency-based spatial weight matrix is defined as^51,52^:

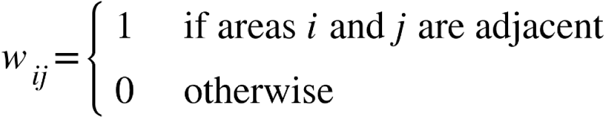

### Spatial Simultaneous Autoregressive Modelling

We first fit an ordinary least squares (OLS) regression model at the univariate and multivariate level for a log-linear distribution of T2D crude rates. The OLS regression equation is expressed as follows:

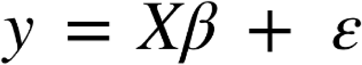

where *y* is a vector of the main outcome variable (i.e., the natural log of T2D rates), *X* is a matrix of observations of the covariates, *β* is a vector coefficient for the covariates, and *ε* is the vector of independent and identically distributed random error terms^51,53^.

We next considered spatial econometric modelling approaches through spatial simultaneous autoregressive (SAR) models (i.e., spatial lag and spatial error models) if the OLS regression residuals exhibited statistically significant spatial autocorrelation, via a Global Moran’s *I* statistic and Lagrange Multiplier tests, using a first-order Queen’s contiguity matrix. A spatial lag model (SLM) examines how logged T2D rates in a district is influenced by the disease burden in adjacent districts. The interpretation of the spatial lag parameter (*ρ*) refers how the average logged T2D rates in neighboring districts is associated with the logged T2D rates of a focal district. The equation is expressed as follows:

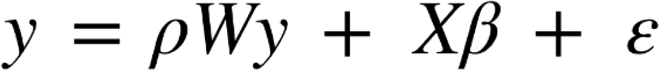

where *y* is the outcome variable (i.e., logged T2D rates), *Wy* is the spatially lagged main outcome variable for y (i.e., the weighted average of outcome variable of neighboring areas), *W* is the spatial weight matrix, *X* is the matrix of observations of the covariates, *β* is a vector coefficient for the covariates, *ε* is the vector of independent and identically distributed random error terms, *ρ* is the spatial autoregressive coefficient for the lagged variable *Wy* and the values range between -1 and 1 [*ρ* > 1 indicates neighborhoods surrounding each other have similar values relative to the outcome (i.e., high or low logged T2D rates); *ρ* < 0 indicates high T2D rates neighborhoods being surrounded by low logged T2D rates neighborhoods and vice versa; *ρ* = 0 indicates no spatial dependence)]^51,53^.

In contrast, a spatial error model (SEM) estimates the extent to which the OLS residual of a district is correlated with its adjacent districts. The spatial error parameter (*λ*) measures the strength of the relationship between the average residuals or errors in neighboring districts and the residual or error of a given district. The SEM equation is expressed as follows:

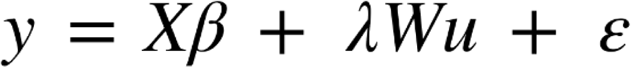

where *y* is the outcome variable (i.e., logged T2D rates), *Wu* is the spatially lagged error term for the main outcome variable *y*, *W* is the spatial weight matrix, *λ* is the spatial autoregressive coefficient for the error terms, *X* is the matrix of observations of the covariates, *β* is a vector coefficient for the covariates, *ε* is the vector of independent and identically distributed random error terms.

### Covariate Selection and Interactions

We note the saturation of our neighborhood covariates that were hypothetically plausible to influence T2D rates by administrative districts. With multiple covariates in the model, there were tendencies for non-spatial and spatial correlations/regression models to be influenced by confounders and interaction effects. We therefore implemented a strategy for covariate selection in the multivariate OLS and spatial models:

i. We first visualized all covariates correlations through scatterplot matrices and assessed these correlations for statistical significance.
ii. We then determined covariates of significant clustering at the univariate spatial autocorrelation level, with positive Global Moran’s *I* values (presence of clustering) as fit to be included in the multivariate OLS and spatial regression models.
iii. We subsequently fit regression models via ordinary least squares (OLS) and assessed potential collinearity between covariates via variation inflation factor (VIF) values. Here, covariates with statistically significant association with the outcome and VIF values of ≤ 5 were determined to be suitable for inclusion in our final multivariate OLS model.
iv. In addition, covariates evidenced to have biological plausibility (i.e. even if the covariate was not statistically significant at the univariate OLS level but importantly being a risk factor for diabetes as evidenced by the national country report, or the literature theoretically, or are biologically influencing predictors like gender, age, or ethnicity) were included in the full multivariate models.

Our approach for regression model building was based on the conceptualization that covariates selected in the models were based on analytical-knowledge thinking (i.e. we considered each covariate alone within the classes then seek to build a set of covariates within each domain of neighborhood covariates, finding the most significant and the least correlated covariates within that domain (visualized in the scatterplot matrices followed by the subsequent steps listed above) Given the grouping of covariates into domains addressing similar types of measures, we avoided generic, automated variable selection techniques (e.g. stepwise approach) that would forcibly select or deselect neighborhood attributes based on statistical significance alone which has a tendency to yield high R-squared values for biased estimates, or cause collinearity within covariates^54^.

We further performed a two-way interaction analysis for the statistically significant attributes from the SAR models to distinguish and quantify the main and interaction effects within the spatial attributes tested in our analysis, an advancement in neighborhood analysis not previously reported elsewhere. All spatial autocorrelations, spatial regressions, and interaction analyses were conducted using GeoDa version 1.18 (Center for Spatial Data Science University of Chicago, IL, USA).

### Interpretations of Statistical Significance and Model Performance

The statistic with the highest value (and lowest *p*-value) will indicate the proper specification for the data. We took a more nuanced approach as suggested recently in the interpretation of statistical significance for models dealing with spatial dependence^53^; that if *p*-value is less than 0.001 (***), between 0.001 and 0.01 (**), between 0.01 and 0.05 (*), between 0.05 and 0.10, or greater than 0.10 to consider the evidence linking main outcome and neighborhood attributes (i.e., against the null hypotheses) to be very strong, strong, moderate, weak, or none, respectively. If spatial models were necessary, the OLS and SAR models were compared using the Akaike Information Criterion (AIC), whereby a lower AIC value indicates a better model fit.

## Results

### Neighborhood Characteristics

Table (1) enumerates neighborhood characteristics by administrative districts in Malaysia. The average proportion of women in a district was 50% while the average proportion of ethnic minorities (Indians) in a district was 4%, with the maximum of 20% Indians being populated in the district of Port Dickson. The average proportion of the majority ethnic group (Bumiputera) in a district was 81%. Regarding age composition, on average, 30% were aged 35-49 years, 21% were aged 50-64 years, and 11% were aged 65 years or older. In an average Malaysian district, the proportion of households living below fifty percent of median income was 16.78%, the proportion of households living below the national poverty line was 10.96%, while the average income inequality and unemployment rates by districts was 0.36 and 5.43% respectively. With rapid urbanization across Malaysia, on average, urban growth rate was 3.18%. For neighborhood safety, the average CCTV coverage per 1000 population was 0.22, with the maximum coverage of 7.85 CCTV per 1000 population in the city of Putrajaya. The average proportion of property crimes per 1000 population was 2.60, while an average of 1.42 per 1000 population using illegal drugs in any neighborhood. The average proportion of population not engaged in fitness, exercise, or sports activities was 44%. The average percentage of areas with access to parks was 68.31%, while the average percentages with access to jogging track, cycling track, mini stadium, football field, gymnasium, and bowling center were 63.92%, 57.74%, 51.57%, 83.64%, 59.8%, and 26.8% respectively.

**Table 1.**
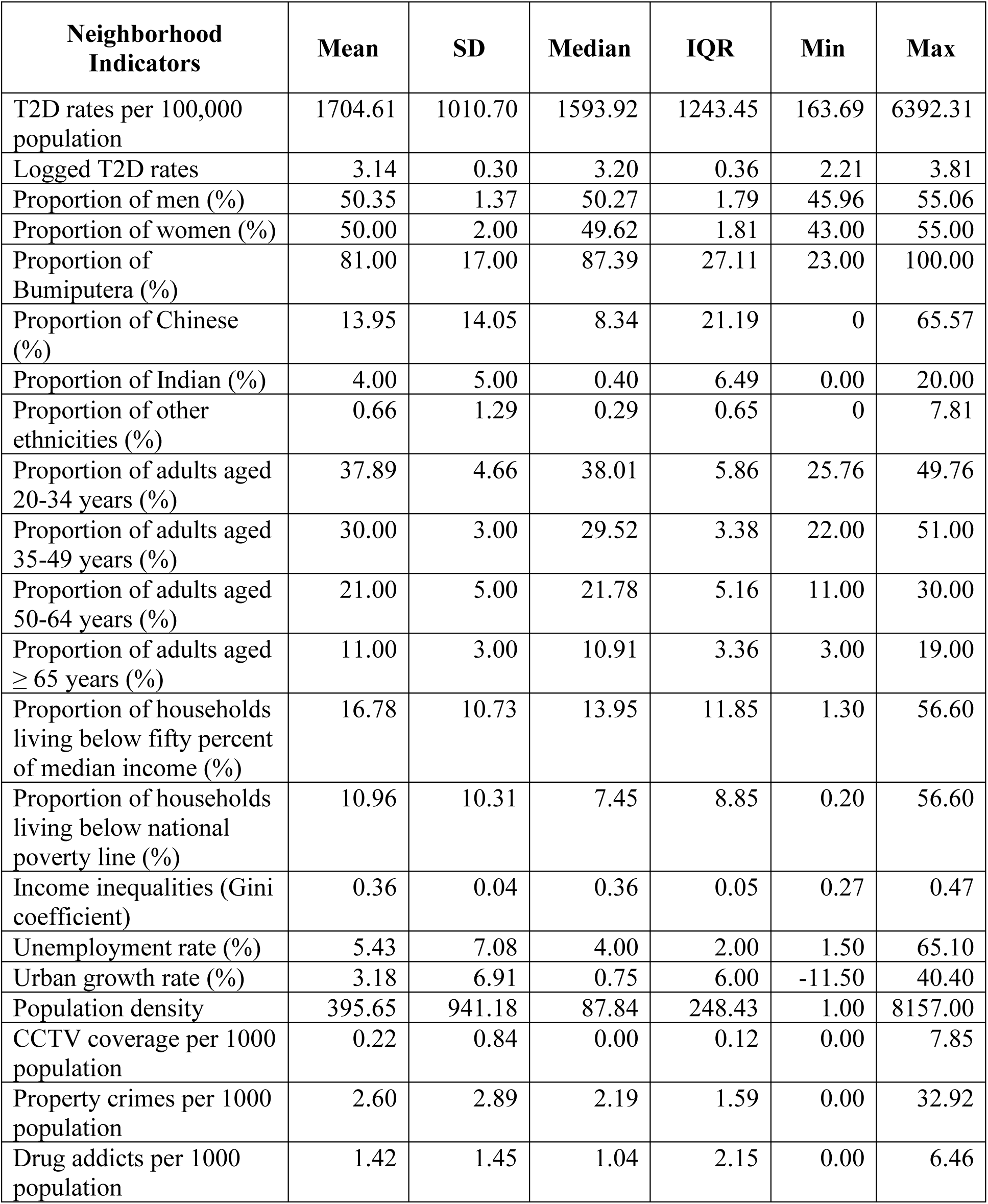
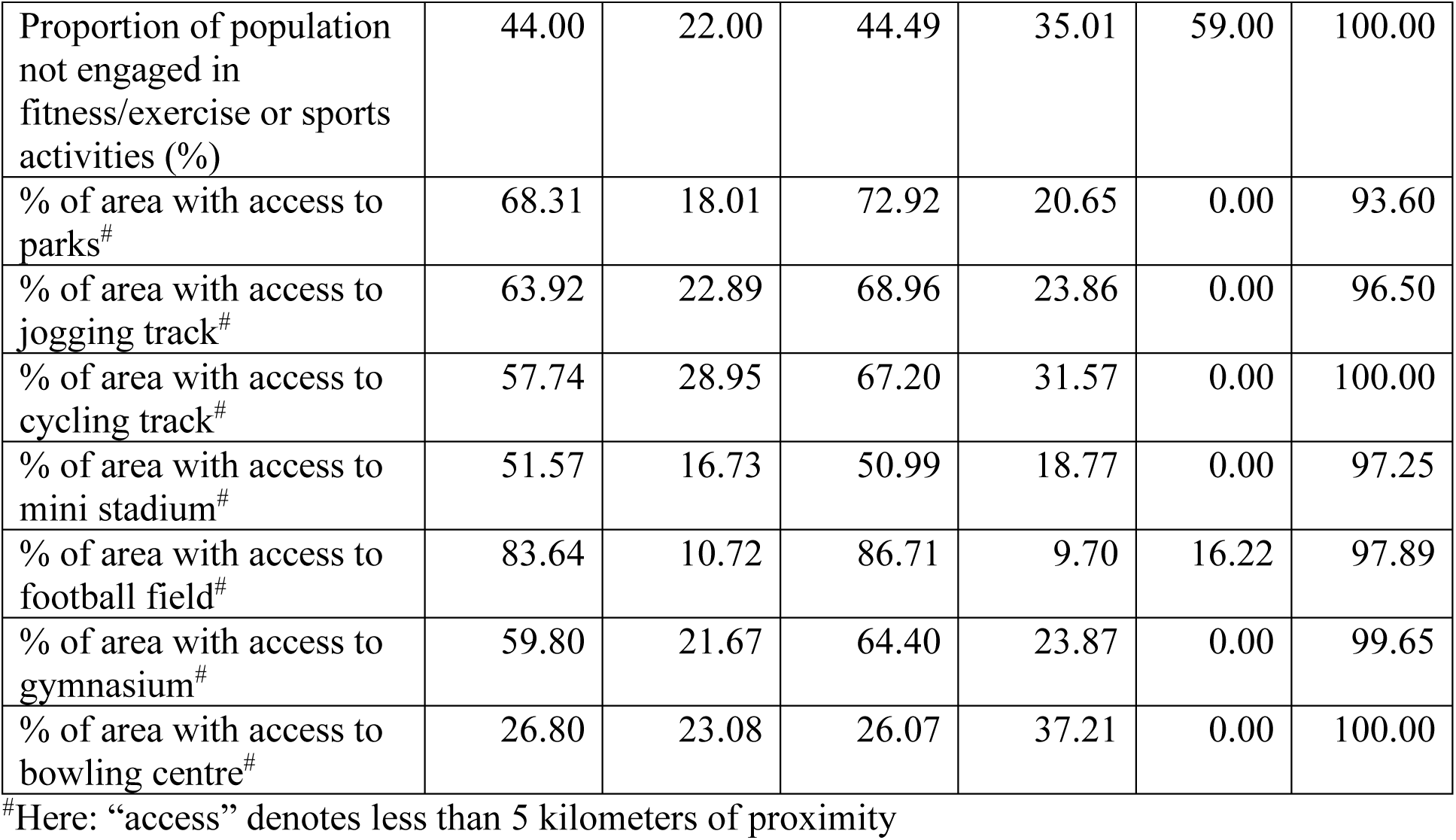
Neighborhood characteristics (n = 144)

### Distribution of Type 2 Diabetes

Because QQ-plots suggests that the distribution of T2D rates was not Gaussian, we log transformed this variable to meet the assumption of normal error distribution for fitting linear regression, spatial autocorrelations, and spatial simultaneous autoregressive (SAR) models (Supplementary Figure 1).

### Logged Transformed Distribution

Figure (1) shows a quantile map of the distribution of logged T2D crude-rates among adults aged ≥20 years by administrative districts in Malaysia. Higher rates of T2D were clustered along the districts nested within the Southern, Central, Northern, and parts of the East Coast regions in the country. For East Malaysia (Borneo) states, T2D rates were highly clustered along the Northwest districts of Sarawak. Lower rates of T2D were mostly clustered in the East Coast region, Northeast region in the state of Sarawak, and most districts in the state of Sabah. Quantile map of the untransformed crude rates is available in Supplementary Figure 2.

**Figure 1.**
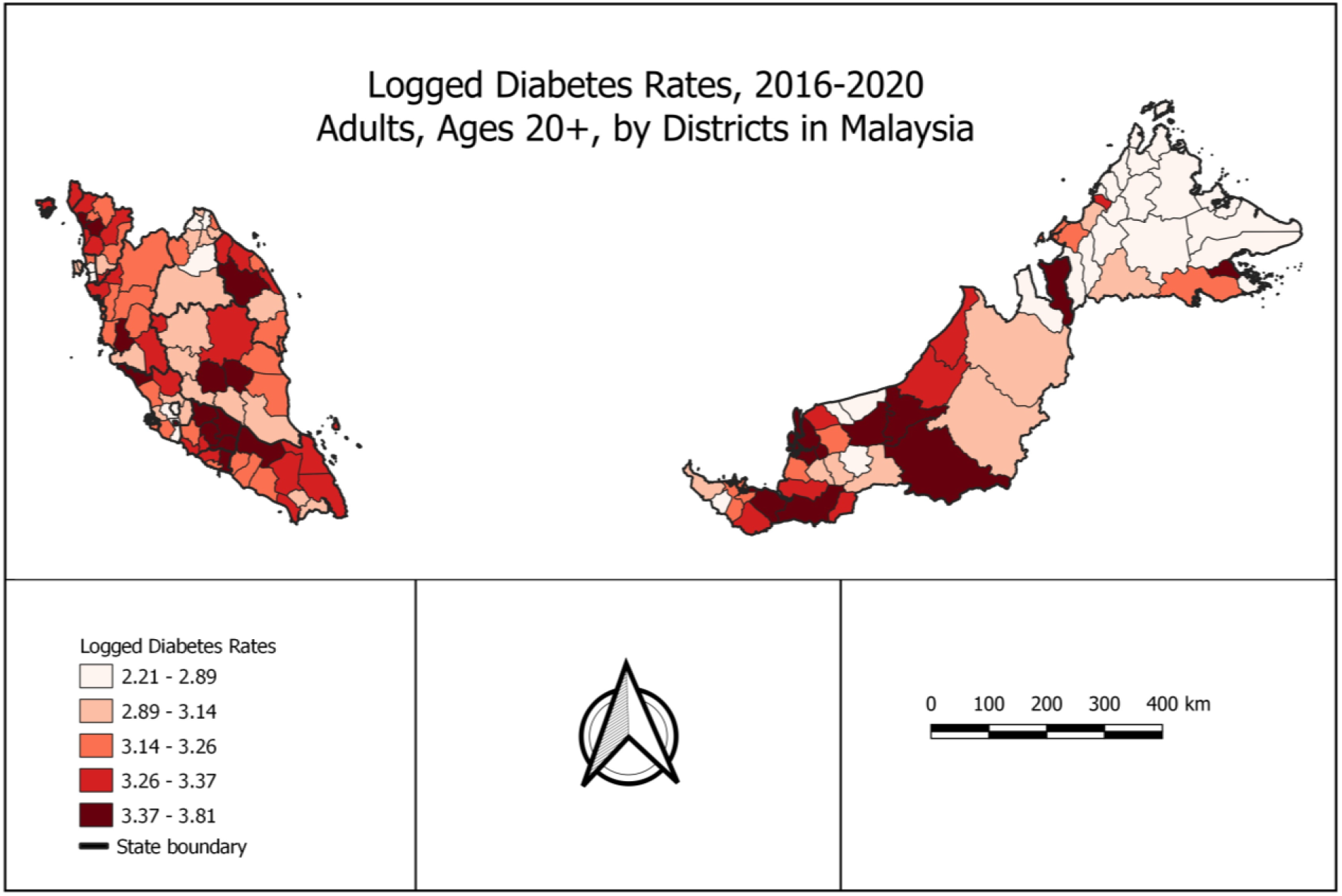
Distribution of T2D rates (logged) by administrative districts in Malaysia

### Geostatistical Interpretation of T2D Distribution

The box-map revealed three upper outliers, namely the districts of Meradong, Sarikei, and Simunjan in the state of Sarawak, but no unusual low-rate outliers. The population mean [standard deviation (SD)] of T2D rates was 1704.61 (1010.70) per 100,000 people, and the rates ranged between 163.70 to 6392.31 per 100,000 population throughout districts in Malaysia. The median [interquartile range (IQR)] of T2D rates distribution at the countrywide level was 1593.92 (1243.45) per 100,000 people. Up to a quarter of districts had less than 972.15 per 100,000 people with T2D; these lower-rate regions were clustered around the North of Kelantan, parts of Selangor, Kedah, North-West Sarawak, and almost all districts in Sabah. A quarter up to half of the districts reported T2D rates to range between 972.15 per 100,000 people and less than 1593.92 per 100,000 people; most of these moderate-rate districts were dense in the state of Pahang, East and West of Sarawak. From half up to three quarter of the districts had T2D rates ranged between 1593.92 per 100,000 people to less than 2215.59 per 100,000 people; these districts were distributed across the Southern, Central, and Northern regions, with some scattered in the East Coast of Malaysia. Over three quarters of the districts had T2D rates ranging between 2215.59 per 100,000 people to less than 4080.76 per 100,000 people; these areas mostly included districts in the Southern, Central, Northern and East-Coast regions. The districts in the North, South, North-West, and South-West areas of Sarawak observed similar crude rates. The visualization of the geographic distribution of crude rates appears in Supplementary Figure 3.

### Rate Smoothing

Since local rates are based on different population sizes, local rate estimates can vary in precision. Empirical Bayes smoothing approaches allow us to pool information from neighboring districts to improve precision for rate estimates from low-population-size districts^52^. The Empirical Bayes smoothed rate map for district-level T2D rates appears in Supplementary Figure 4. In this application, the observed distribution of smoothed rates does not vary much from that of the logged transformed crude rates, due to the relatively high rate of T2D compared to the rarer health outcomes (e.g., cancer) often featured in small area estimates of local rates.

### Non-Spatial and Spatial Correlations of Diabetes Rates

The Global Moran’s *I* (spatial autocorrelation) for logged T2D rates was 0.370 (*p* = 0.001), indicating a significant positive spatial autocorrelation. The Local Indicators of Spatial Autocorrelation (LISA), i.e., the local Moran’s index, identified spatial clusters where high rates occurred near other high rates across districts within the state of Negeri Sembilan, Kedah, and parts of Sarawak, whereas clusters of low rates near other low rates were concentrated across rural districts in Sabah and small areal districts in Selangor (Figure 2A). Figure (2B) enumerates the LISA significance of these clusters.

**Figure 2.**
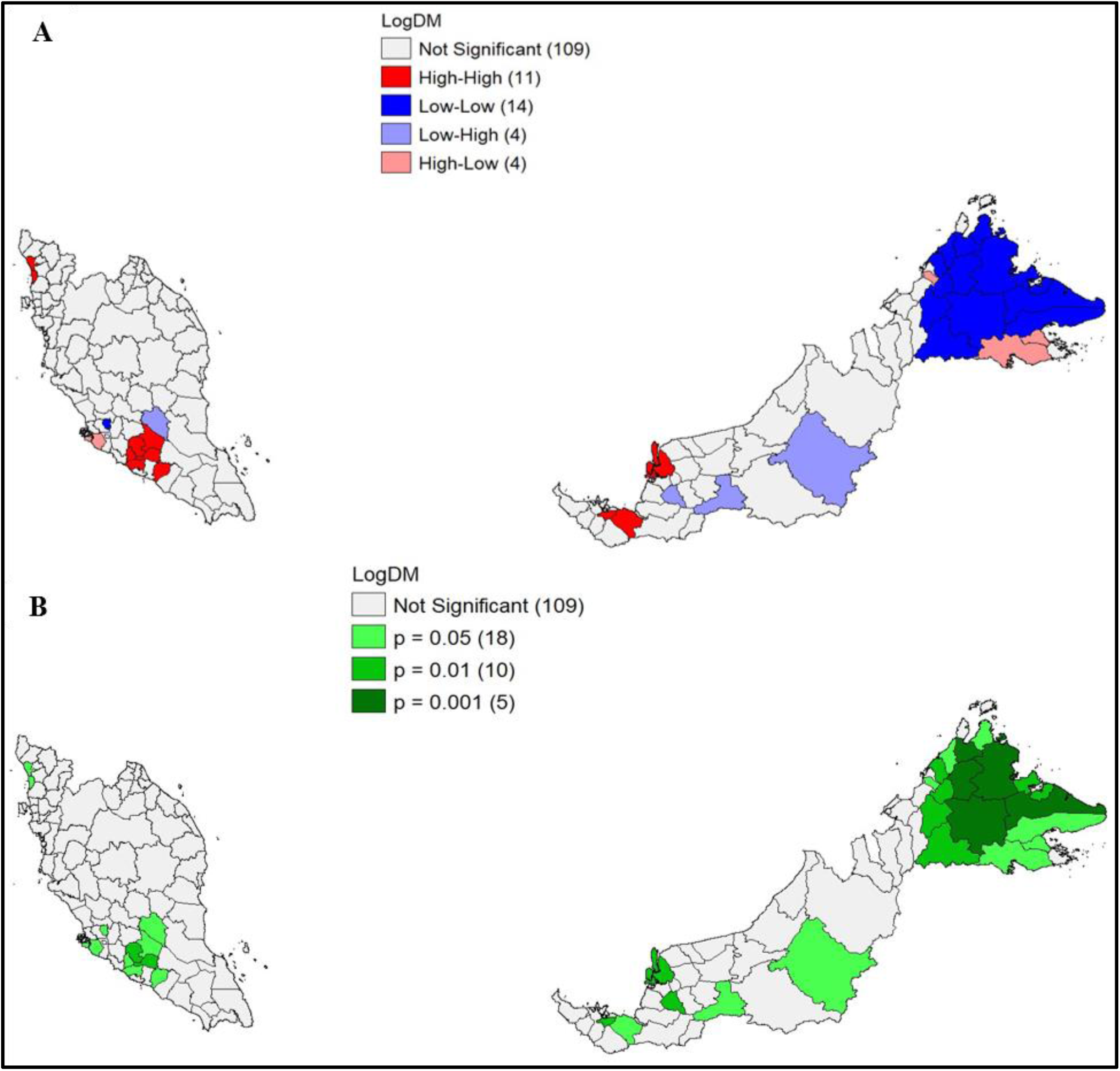
[A] Univariate LISA cluster map; [B] Univariate LISA significance map of spatial clustering and outliers

The Global Moran’s *I* for neighborhood characteristics appears in Table (2). The spatial autocorrelation analysis (*I*) had eleven additional statistically significant coefficients in contrast to the statistically significant coefficients available in the non-spatial conventional linear correlations (*r*). We found that the proportion of women, the proportion of Bumiputera, the proportion of Indians, the proportion of adults aged 35-49 years, 50-64 years, and 65 years or older, proportion of households living below 50% of median income, proportion of households living below national poverty line, income inequalities, drug users per 1000 population, proportion of population not engaged in fitness/exercise or sports activities, percentage of areas with access to parks, jogging track, cycling track, football field, gymnasium, and bowling center within five kilometers of proximity from neighborhoods showed spatial autocorrelation coefficients larger than conventional linear correlation coefficients (*I* > *r*), suggesting that these correlations were highly determined by geographic locations.

**Table 2.**
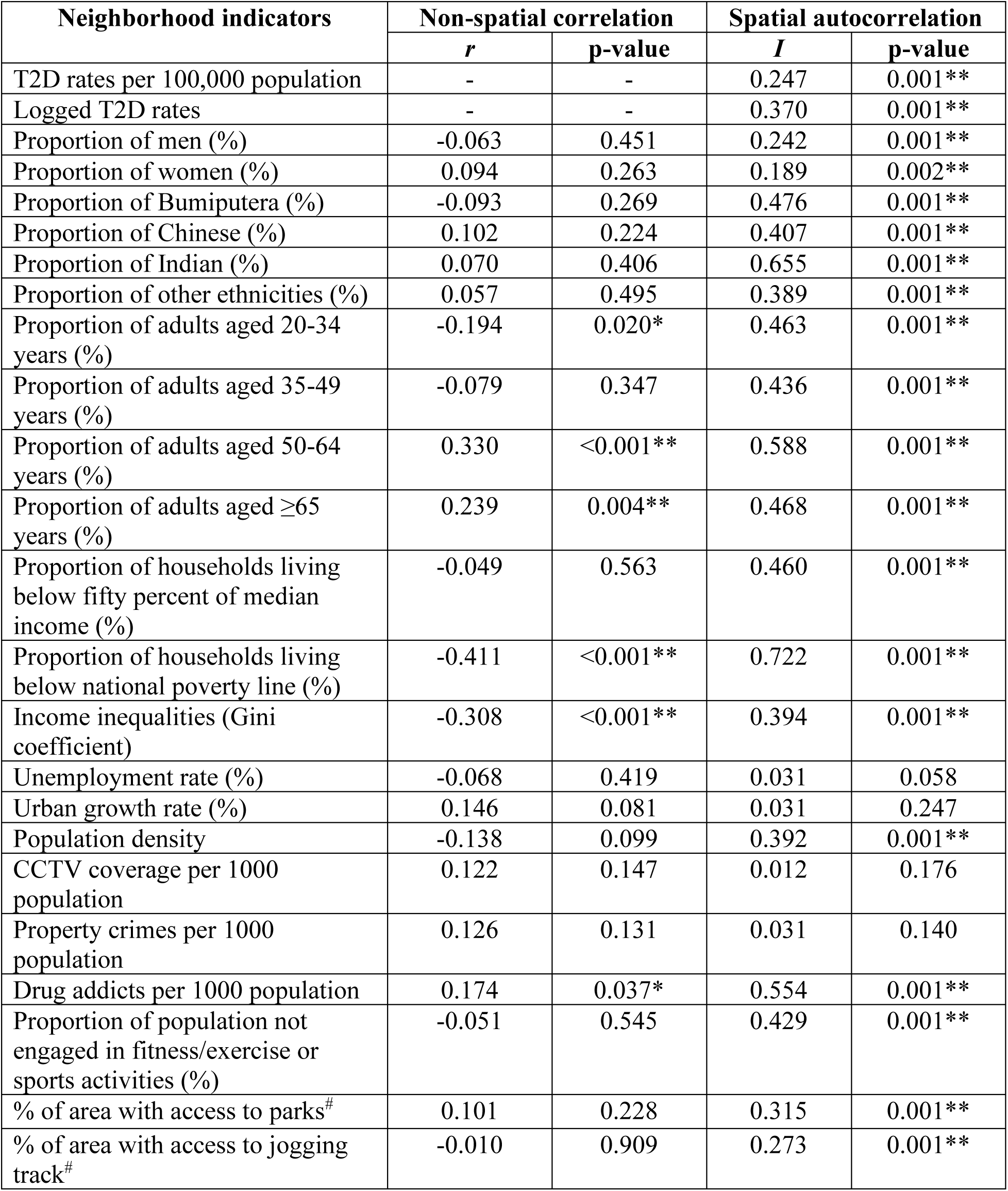
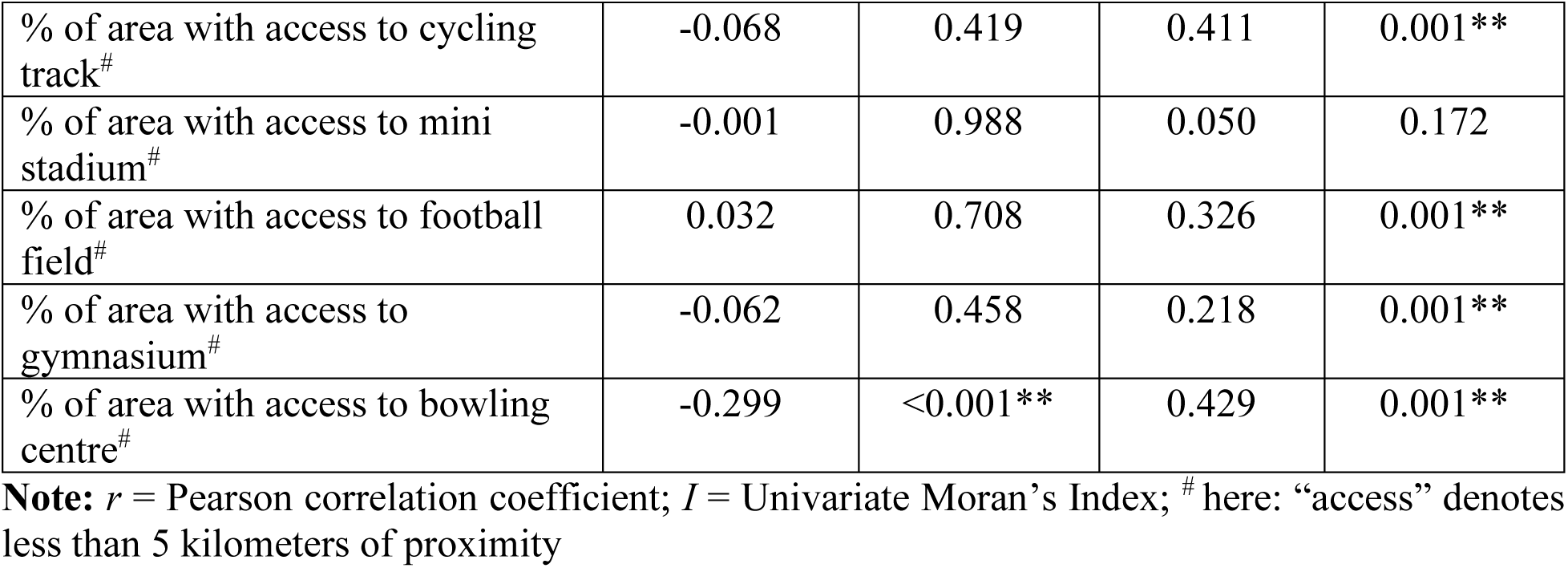
Non-spatial correlation with logged T2D rates and spatial autocorrelations for covariates reported from small areas (n = 144)

| Neighborhood indicators | Non-spatial correlation |  | Spatial autocorrelation |  |
| --- | --- | --- | --- | --- |
|  | *r* | p-value | *I* | p-value |
| T2D rates per 100,000 population | - | - | 0.247 | 0.001** |
| Logged T2D rates | - | - | 0.370 | 0.001** |
| Proportion of men (%) | -0.063 | 0.451 | 0.242 | 0.001** |
| Proportion of women (%) | 0.094 | 0.263 | 0.189 | 0.002** |
| Proportion of Bumiputera (%) | -0.093 | 0.269 | 0.476 | 0.001** |
| Proportion of Chinese (%) | 0.102 | 0.224 | 0.407 | 0.001** |
| Proportion of Indian (%) | 0.070 | 0.406 | 0.655 | 0.001** |
| Proportion of other ethnicities (%) | 0.057 | 0.495 | 0.389 | 0.001** |
| Proportion of adults aged 20-34 years (%) | -0.194 | 0.020* | 0.463 | 0.001** |
| Proportion of adults aged 35-49 years (%) | -0.079 | 0.347 | 0.436 | 0.001** |
| Proportion of adults aged 50-64 years (%) | 0.330 | <0.001** | 0.588 | 0.001** |
| Proportion of adults aged ≥65 years (%) | 0.239 | 0.004** | 0.468 | 0.001** |
| Proportion of households living below fifty percent of median income (%) | -0.049 | 0.563 | 0.460 | 0.001** |
| Proportion of households living below national poverty line (%) | -0.411 | <0.001** | 0.722 | 0.001** |
| Income inequalities (Gini coefficient) | -0.308 | <0.001** | 0.394 | 0.001** |
| Unemployment rate (%) | -0.068 | 0.419 | 0.031 | 0.058 |
| Urban growth rate (%) | 0.146 | 0.081 | 0.031 | 0.247 |
| Population density | -0.138 | 0.099 | 0.392 | 0.001** |
| CCTV coverage per 1000 population | 0.122 | 0.147 | 0.012 | 0.176 |
| Property crimes per 1000 population | 0.126 | 0.131 | 0.031 | 0.140 |
| Drug addicts per 1000 population | 0.174 | 0.037* | 0.554 | 0.001** |
| Proportion of population not engaged in fitness/exercise or sports activities (%) | -0.051 | 0.545 | 0.429 | 0.001** |
| % of area with access to parks<sup>#</sup> | 0.101 | 0.228 | 0.315 | 0.001** |
| % of area with access to jogging track<sup>#</sup> | -0.010 | 0.909 | 0.273 | 0.001** |
| % of area with access to cycling track<sup>#</sup> | -0.068 | 0.419 | 0.411 | 0.001** |
| % of area with access to mini stadium<sup>#</sup> | -0.001 | 0.988 | 0.050 | 0.172 |
| % of area with access to football field<sup>#</sup> | 0.032 | 0.708 | 0.326 | 0.001** |
| % of area with access to gymnasium<sup>#</sup> | -0.062 | 0.458 | 0.218 | 0.001** |
| % of area with access to bowling centre<sup>#</sup> | -0.299 | <0.001** | 0.429 | 0.001** |

**Note:** $r$ = Pearson correlation coefficient; $I$ = Univariate Moran's Index; <sup>#</sup> here: "access" denotes less than 5 kilometers of proximity

### Baseline Regressions

We synthesized a baseline univariate ordinary least squares (OLS) regression with VIF values to determine covariates to be included in the multivariate OLS and SAR models (Supplementary Table 1). All covariates with statistical significance, VIF values ≤ 5, significant clustering, and biologically plausible were included in the full multivariate OLS model (Table 3). Subsequently we evaluated the diagnostic performance of the model if further subjected for spatially weighted spatial lag or error models.

**Table 3.**
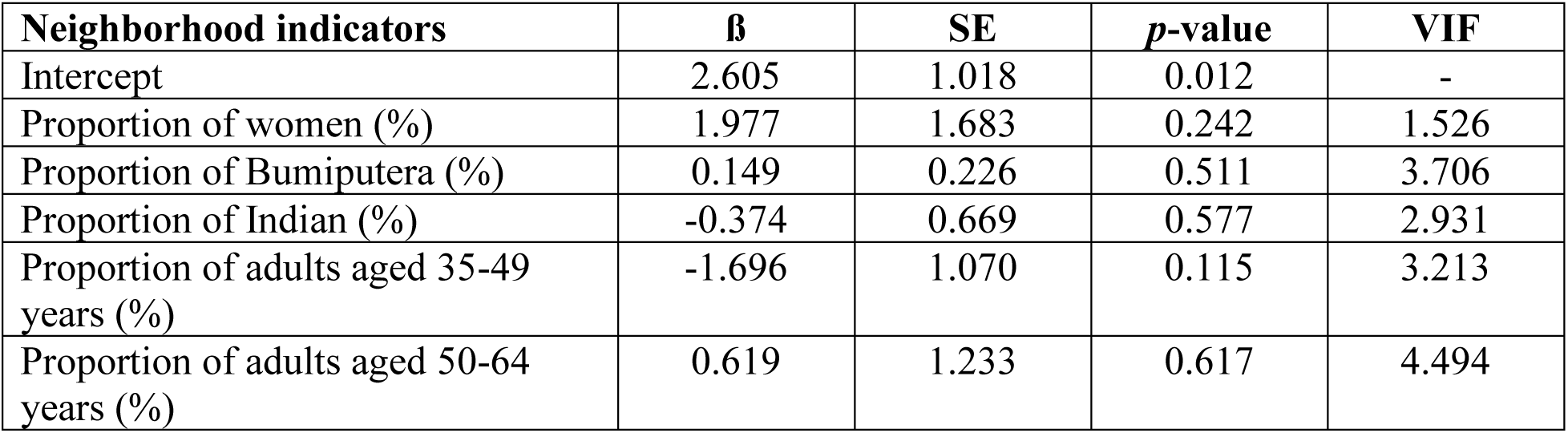
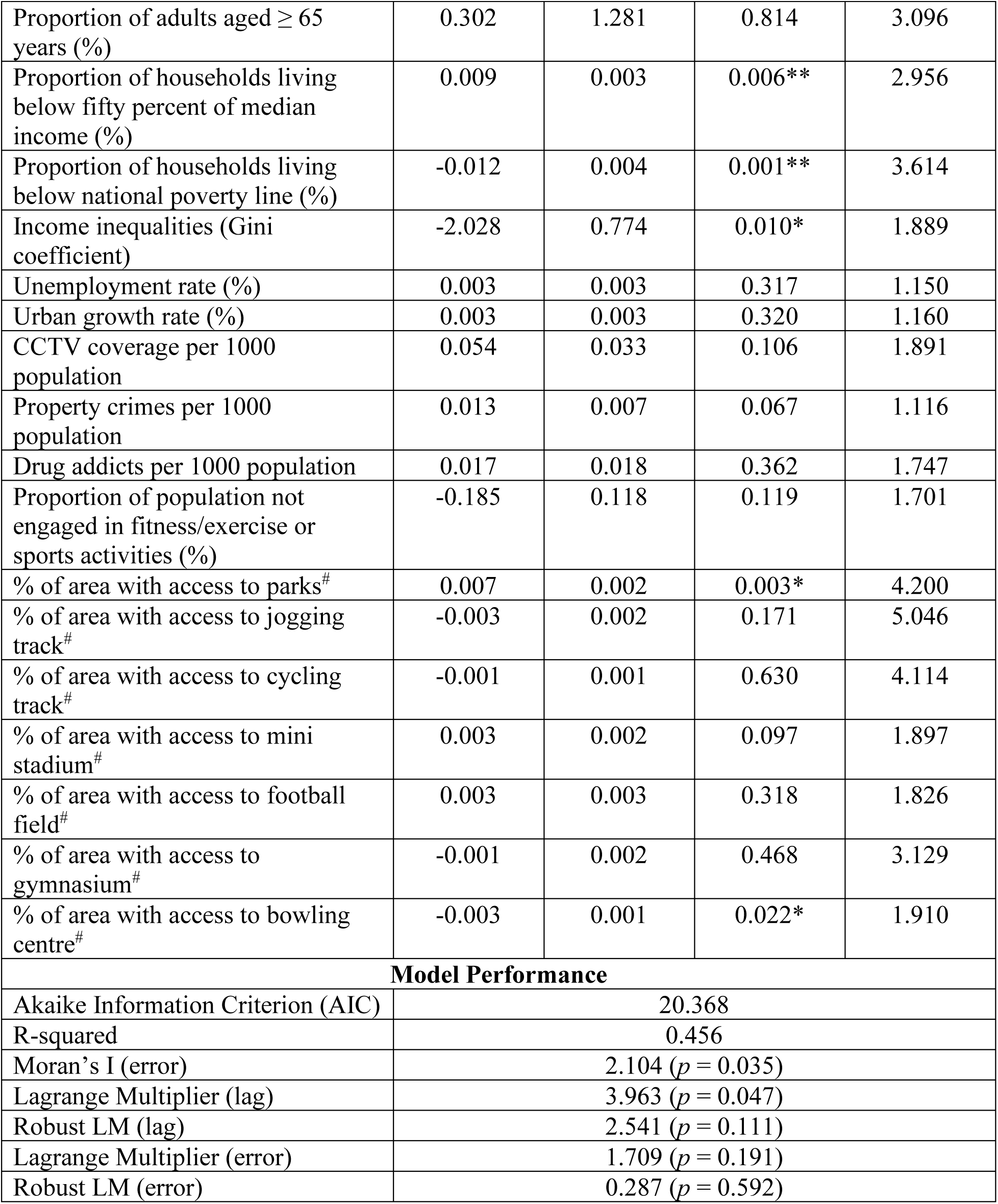
Full multivariate OLS model estimation on the association between neighborhood indicators and local logged diabetes rates (n = 144)

### Full Multivariate Ordinary Least Squares (OLS) Model Estimation on the Association between Neighborhood Indicators and Logged T2D Rates

Table (3) exhibits the findings of the full multivariate ordinary least squares (OLS) model estimation on the associations between neighborhood indicators and T2D rates (logged). On the influence of neighborhood’s socio-economic vulnerability, there was strong evidence that T2D rates had a positive relationship with the proportion of households living below fifty percent of the median income; each one percent increase of households living below fifty percent of the median income was associated with 0.009 increase of T2D rates (β = 0.009, *p* = 0.006). However, there was strong evidence that T2D rates had a negative relationship with the proportion of households living below the national poverty line; each one percent increase of households living below the national poverty line was associated with 0.012 decline of T2D rates (β = 0.009, *p* = 0.006). There was moderate evidence that T2D rates had a negative relationship with income inequalities; each one unit rise in income inequalities was associated with 2.028 decline of T2D rates (β = -2.028, *p* = 0.010).

For neighborhood’s accessibility to fitness facilities, there was strong evidence that T2D rates had a positive relationship with areal proximity to parks; each additional one percent of areal proximity to a park was associated with 0.007 percent rise of T2D rates (β = 0.007, *p* = 0.003). In contrast, there was moderate evidence that T2D rates had a negative relationship areal proximity to a bowling center; each additional one percent of areal proximity to a bowling center was associated with a 0.003 decline of T2D rates (β = - 0.003, *p* = 0.022) (Table 3).

The OLS model also weakly adjusted for one neighborhood safety covariate (i.e., property crimes per 1000 population, β = 0.013, *p* = 0.067) and one neighborhood accessibility to fitness facility covariate (i.e., access to mini stadium, β = 0.003, *p* = 0.097). A standard deviation map of residuals from the OLS regression is exhibited in Supplementary Figure 5. The p-value for the Global Moran’s *I* residuals of the OLS model was statistically significant (*p* = 0.035). Similarly, the p-values for the Lagrange Multiplier tests (lag value) were statistically significant, indicating that a subsequent execution of the spatial lag model (SLM) was necessary (Table 3). The model accounted for 45.6% of the total variance for the relationships between T2D rates and neighborhood attributes.

### Full Multivariate Spatial Lag Model (SLM) Estimation on the Association between Neighborhood Indicators and Logged T2D Rates

Table (4) shows the results of the spatial lag model (SLM) estimation on the associations between neighborhood indicators and T2D rates (logged). Overall, the coefficients of the covariates in the SLM model were further adjusted than those yielded in the OLS model. The statistical significance of the coefficients was stronger in the SLM model as compared to the OLS model, with an addition of two covariates: CCTV coverage and property crimes having moderate strength of significance in the model (*p* = 0.023 and *p* = 0.033 respectively).

**Table 4.** Full multivariate SLM model on the association between neighborhood indicators and logged diabetes rates (n = 144)

| Neighborhood indicators | $\beta$ | SE | $p$-value |
| --- | --- | --- | --- |
| Intercept | 2.712 | 0.923 | 0.003 |
| Proportion of women | 1.497 | 1.539 | 0.331 |
| Proportion of Bumiputera | 0.131 | 0.205 | 0.521 |
| Proportion of Indian | -0.394 | 0.604 | 0.515 |
| Proportion of adults aged 35-49 years | -1.863 | 0.974 | 0.056 |
| Proportion of adults aged 50-64 years | 0.321 | 1.126 | 0.775 |
| Proportion of adults aged $\geq 65$ years | 0.206 | 1.158 | 0.859 |
| Proportion of households living below fifty percent of median income (%) | 0.009 | 0.003 | 0.002** |
| Proportion of households living below national poverty line (%) | -0.012 | 0.003 | 0.001** |
| Income inequalities (Gini coefficient) | -2.005 | 0.699 | 0.004** |
| Unemployment rate (%) | 0.004 | 0.003 | 0.177 |
| Urban growth rate (%) | 0.004 | 0.003 | 0.177 |
| CCTV coverage per 1000 population | 0.070 | 0.031 | 0.023* |
| Property crimes per 1000 population | 0.014 | 0.007 | 0.033* |
| Drug addicts per 1000 population | 0.022 | 0.017 | 0.185 |
| Proportion of population not engaged in fitness/exercise or sports activities (%) | -0.173 | 0.107 | 0.106 |
| % of area with access to parks<sup>#</sup> | 0.007 | 0.002 | 0.001** |
| % of area with access to jogging track<sup>#</sup> | -0.002 | 0.002 | 0.170 |
| % of area with access to cycling track<sup>#</sup> | -0.001 | 0.001 | 0.473 |
| % of area with access to mini stadium<sup>#</sup> | 0.003 | 0.002 | 0.051 |
| % of area with access to football field<sup>#</sup> | 0.002 | 0.002 | 0.393 |
| % of area with access to gymnasium<sup>#</sup> | -0.001 | 0.001 | 0.377 |
| % of area with access to bowling centre<sup>#</sup> | -0.003 | 0.001 | 0.019* |
| **Model Performance** |  |  |  |
| Akaike Information Criterion (AIC) | 18.403 |  |  |
| R-squared | 0.472 |  |  |
| Lag Coefficient ($\rho$) | 0.048 | | |
| Likelihood Ratio Test | 3.965 (p = 0.046) |  |  |

With reference to neighborhood socio-economic vulnerability, each additional one percent of households living below fifty percent of the median income was associated with 0.009 percent rise of logged T2D rates, with all other covariates held constant (β = 0.009, *p* = 0.002). In contrast, each additional one percent of households living below the national poverty line was associated with 0.012 percent decline of logged T2D rates, with all other covariates held constant (β = - 0.012, *p* = 0.001). Each additional one unit rise of income inequality was associated with 2.005 percent decline of logged T2D rates, with all other covariates held constant (β = - 2.005, *p* = 0.004) (Table 4).

On the influence of neighborhood safety, we found that each additional one unit rise of CCTV coverage was associated with 0.070 percent increase of logged T2D rates, with all other covariates held constant (β = 0.070, *p* = 0.023). Each additional one unit rise in property crimes was associated with 0.014 percent increase of T2D rates, with all other covariates held constant (β = 0.014, *p* = 0.033) (Table 4).

For neighborhood’s accessibility of built-environments, we found that each additional one percent rise of areal proximity to a park was associated with a 0.007 percent increase of logged T2D rates (β = 0.007, *p* = 0.001), and each additional one percent rise of areal proximity to a bowling center was associated with a 0.003 percent decline of logged T2D rates ((β = - 0.003, *p* = 0.019) (Table 4).

The SLM model also weakly adjusted for one neighborhood demography covariate (i.e., proportion of adults aged 35-49 years, *p* = 0.056) and one neighborhood accessibility to fitness facility covariate (i.e., mini stadium, *p* = 0.051) (Table 4). The model accounted for 47.2% of the total variance for the relationships between logged T2D rates and neighborhood attributes. Residuals are reported in Supplementary Figure 6.

### Evaluating Two-Way Interactions between Spatial Attributes

The geo-visualization of bivariate quantile maps for the statistically significant covariates in the SLM model that influenced the distribution of T2D rates by administrative districts is available in Supplementary Figures 7-13.

Next, we ran 31 two-way interaction modelling analyses contextualized within the framework of statistical significance from the full multivariate SLM model to investigate the interactions between (1) access to fitness facilities with neighborhood poverty and inequity; (2) access to fitness facilities with neighborhood safety and inequity; and (3) access to fitness facilities with neighborhood poverty and safety on the influence T2D. These models allow logical interpretations within the SLM model exploring attenuating or accelerating influences of interaction effects influencing neighborhood T2D rates in Malaysia.

In the SLM two-way interaction modelling analysis that evaluated access to fitness facilities with neighborhood poverty and inequity, we found significant negative association with local income inequality (β = -6.952, p = 0.002) but no significant association with the proportion of households living below 50% of median income (β = 0.004, p = 0.134). The effect of local income inequality was moderated by the presence of parks as indicated by the significant interaction for (parks*income inequality) (β = 0.063, p-value for interaction = 0.037) (Model 3, Supplementary Table 2).

In assessing two-way interactions between access to fitness facilities with neighborhood safety and income inequity covariates, the SLM model showed a relatively weak significance for interaction terms in the SLM model: (parks*income inequalities) (β = 0.059, p-value = 0.047), and no significant interaction with local CCTV coverage per 1000 population (β = 0.048, p = 0.069) (Model 4, Supplementary Table 3).

Finally, we assessed the two-way interactions between access to fitness facilities with neighborhood poverty and safety. We found significant positive associations with the main effects, property crimes per 1000 population (β = 0.060, p = 0.005) and access to parks (β = 0.005, p = 0.003), but negative associations with the proportion of population living below poverty line (β = -0.011, p<0.001) or access to bowling center (β = -0.005, p<0.001). However, the interaction effect (parks*property crimes) revealed significant negatively association with T2D distribution (β = - 0.001, p-value = 0.010) (Model 5, Supplementary Table 4).

Taken together, we find significant local associations between local measures of income inequality, property crimes per 1000 population, and access to parks. However, these variables, and other measures of local physical activity locations, also reveal moderated effects on T2D in the presence of other local measures of income, access to physical activity, and safety. These interactions suggest further research exploring the specific neighborhood compositions of these features to provide insight on local prevention measures.

## Discussion

We found that areas with high T2D rates were predominantly concentrated along the West of Peninsular Malaysia, across districts in the Southern, Central, and Northern regions, and parts of the areal districts in the North, South, and West of Sarawak state. We observed significant spatial autocorrelations on T2D rates and neighborhood indicators, suggesting that these areal-level indicators had fully or partly influenced the formation of T2D clusters across local geographies in Malaysia.

As noted above, our final SLM model revealed a triad of interactions between neighborhood socio-economic vulnerabilities, neighborhood safety features, the influence of local residential access to neighborhood built-environments fitness activities, and T2D burden. These associations were consistent with previous geospatial investigations from the USA^14-16,55^, Saudi Arabia^56^, Australia^57^, Bangladesh^58^, and Brazil^59^. We note that these studies originate from both developed and developing nations with different geo-political, social, and cultural norms; alongside differences of spatial attributes within the rural-urban matrix. The applications of place-based correlations and regression modelling at different spatial scales in those studies, either at the regional, state, or county levels yielded different directions of relationships of the effect sizes when linking with local neighborhood risks, a condition susceptible to the modifiable areal unit problem (MAUP) and provide insight into the spatial scale of main effects and their interactions on local risk. Our study explored neighborhood effects at the district level in Malaysia, a boundary used by the national population census estimates that allow smaller area estimates for policy making.

Income and poverty are strong population health indicators with higher income typically indicative for better health. Studies have found positive correlations between these indicators and T2D^60,61^, but our study found otherwise. A unit rise in the proportion of households living below the national poverty line was related to a decrease in T2D rates, but a unit rise in the proportion of households living below fifty percent of the median household income increased the risk of T2D across neighborhoods. We note that the metrics of spatial economic covariates through spatially adjusted econometric approaches with ecological linkages to T2D rates have different cut-offs and that the income distributions in Malaysia have local associations with the urban/rural geography of the country. Specifically, the proportion of households living below the national poverty line (i.e., extreme poor with monthly income cut-off of USD 533.10) were mostly distributed across the rural districts in the state of Sabah, and parts of the rural districts within the East Coast region of the country – these areas had low T2D rates (see joint distributions in Supplementary Figures 7-13). In contrast, the proportion of households living below fifty percent of median household income (i.e., households with income cut-off of USD 709.54) had positive relationships with T2D rates. Although not within the extremes of poverty, this income group exists within the upper end of the bottom 40% (B40) lower income group classification in Malaysia that were densely distributed in most rural districts, with some clusters observed across the urban districts in the country. The distribution of T2D rates within this income group was mostly dense across the districts in East Malaysia and Southern regions, with some clusters around the Northern and Central metropolitans, and townships within the East Coast regions (see joint distributions in Supplementary Figures 7-13), consistent with the effect direction of our regression models. Inconsistencies of spatial distribution between different economic covariates linking to T2D or obesity rates persisted in previous works^8,62-67^, suggesting that local interactions of income with other risk factors may moderate observed associations. Other plausible explanations for differing associations could be that extreme poor communities in rural areas have limited health literacy or access to health facilities for T2D screening, whereas the urban poor had better access to subsidized and affordable health services for T2D screening, again, suggesting interactions between risk factors at the neighborhood level.

While poverty accelerates disease risk and poor health outcomes, the associations between income inequality as measured by the Gini coefficient exhibited inconsistencies throughout the spatial epidemiology literature^8,68,69^. We found that a unit rise in income inequality decreased T2D rates, when principally it was anticipated to be high, a condition most likely attributable to the “Swiss paradox” wherein increased income inequality that supposedly cause negative health impacts would inversely show better population health outcomes when measured on a finer geographical scale of aggregation^68^. These areas mostly comprised of districts within the rural regions of similar population distributions residing below the national poverty line as described earlier. As income inequality is a contextual variable specific to geographical scales influenced by local social or commercial drivers, we note that inconsistencies between the association of income inequality and chronic conditions can be influenced by the different spatial measurement units (e.g., state, district, or county levels) or the scales used (i.e., a single measure through Gini coefficient or composite measures of neighborhood deprivation scores), and, as suggested by our interaction analysis, potential mediators or confounders that change the degree of associations where areal level attributes within neighborhood characteristics interact with local health outcomes^6,8^.

Inequalities and poverty collectively pressure local communities within deprived neighborhoods, causing poor health outcomes. We postulate plausible psychosocial and neo-materialist theories whereby different social gradients within community hierarchy pose different levels of stress within neighborhood living circumstances, in addition to poor communities paying less attention to health needs, practicing healthy lifestyles, or having easy accessibility to health or fitness facilities as compared to richer ones^70-72^. Deprived neighborhoods are often compromised with neighborhood safety features as consequence of social delinquency, accelerated local crime rates, or drug addiction. Our findings showed that apart from poverty attributes, neighborhood safety features interacted with access to fitness facilities (i.e., bowling centers) and parks, likely due to fear and psychosocial stress depriving local communities from actively participating in physical fitness, a circumstance that accelerate incidence of obesity and T2D rates in local neighborhoods, consistent with previous literature^6,8,15,16^.

## Strengths and Limitations

Our countrywide results provide baseline comprehensive evidence for defining local neighborhood policy interventions to control the T2D burden at the district level in Malaysia. We note areal disparities of different neighborhood attributes and clusters of T2D, thereby suggesting local behavioral health interventions, health promotions, or local townships restructure or sustainability efforts based on specific targets and areas rather than the practice of implementing generic interventions nationwide. As policies are implemented at an aggregate level rather than being individualized, the ecological regression approach promptly pointed out areas of high, low, or epidemiologically transitioning areas of local diabetes burden with their associated place-based characteristics, guiding planning, implementation, and ongoing evaluation of local public health responses.

The data used in this study are reliable, accurate, and generalizable, as countrywide disease registries are official health data captured through proper diagnostics by health personnels using validated clinical practice guidelines, thereby tackling many limitations from more commonly used cross-sectional surveys that are subject to non-generalizability, social-desirability bias, compromised accuracies due to measurements used, and potential residual spatial correlation. Our use of spatial modelling approaches accounts for spatial autocorrelations and spillover effects^73-76^, thus strengthening evidence and interpretation of effect associations.

We also acknowledge limitations of the study. While the data were aggregated to the district level in the NDR database, we note that T2D cases were captured based on the primary care clinic’s location in that district’s established catchment area (i.e., principally within five kilometers of residence from the clinic). Therefore, we were unable to establish spatial-analytical connections to the smallest spatial unit (i.e. sub-district or mukim levels) for more robust estimates as they may share local borders or corners that may contaminate the exact location of analysis within the anonymized residence address of the NDR data.

## Conclusion

Our study established that district-level T2D rates in Malaysia are influenced by neighborhood-level socio-economic vulnerability, safety, and access to fitness facilities. The interactions between these covariates provides insight into the patterns of these risk factors, and their interacting effects, at this geographic scale, allowing for localized planning of public health interventions to reduce risk.

## Patient Consent for Publication

Not applicable.

## Ethics Statement

Ethical approval was granted from the ethics committee of Universiti Kebangsaan Malaysia (UKM), Ministry of Higher Education Malaysia (JEP-2022-445) and the Medical Research Ethics Committee (MREC), Ministry of Health Malaysia [NMRR ID-22-01264-EE7 (IIR)].

## Author Contributions

K.G. and M.R.A.M. designed the study. M.R.A.M., N.S., F.I.M. and M.F.M.R. had access and collected data. N.S. and M.F.M.R. managed the data. K.G. and L.A.W. conducted the data analysis. K.G., M.R.A.M., N.S., L.A.W., K.N.A.M. and F.I.M. contributed to the interpretation of the data. K.G. drafted the first version of the manuscript. M.R.A.M., L.A.W. and KNAM critically revised the manuscript for important intellectual content. All authors read and approved the final manuscript.

## Acknowledgement

We thank the various government agencies and ministries in support of this work. We acknowledge the use of Royal Malaysia Police force and National Anti-Drug Agency data by the Ministry of Home Affairs of Malaysia, social population statistics of CCTV coverage data by the Ministry of Housing and Local Government of Malaysia, population census (MyCensus 2020) and Population Wellbeing Fitness Survey data, all of which were sourced through the Department of Statistics Malaysia (DOSM) under the Ministry of Economy Malaysia. We acknowledge the support by the Ministry of Higher Education Malaysia and the Universiti Kebangsaan Malaysia for funding this work and the resources provided to execute this countrywide study. We also thank the Non-Communicable Disease Section, Public Health Division of the Ministry of Health Malaysia, and the Office of Director General of Public Health for providing the National Diabetes Register (NDR) data and the support of this study to be in line with Malaysia’s Public Health Priorities target.

## Data Availability Statement

The data that support the findings of this study are available from the authors but restrictions apply to the availability of these data, which were used under license from the relevant agencies for the current study, and so are not publicly available. Data are, however, available from the authors upon reasonable request and with permission from the relevant agencies or ministries.

---

**Supplementary Figure 1.**
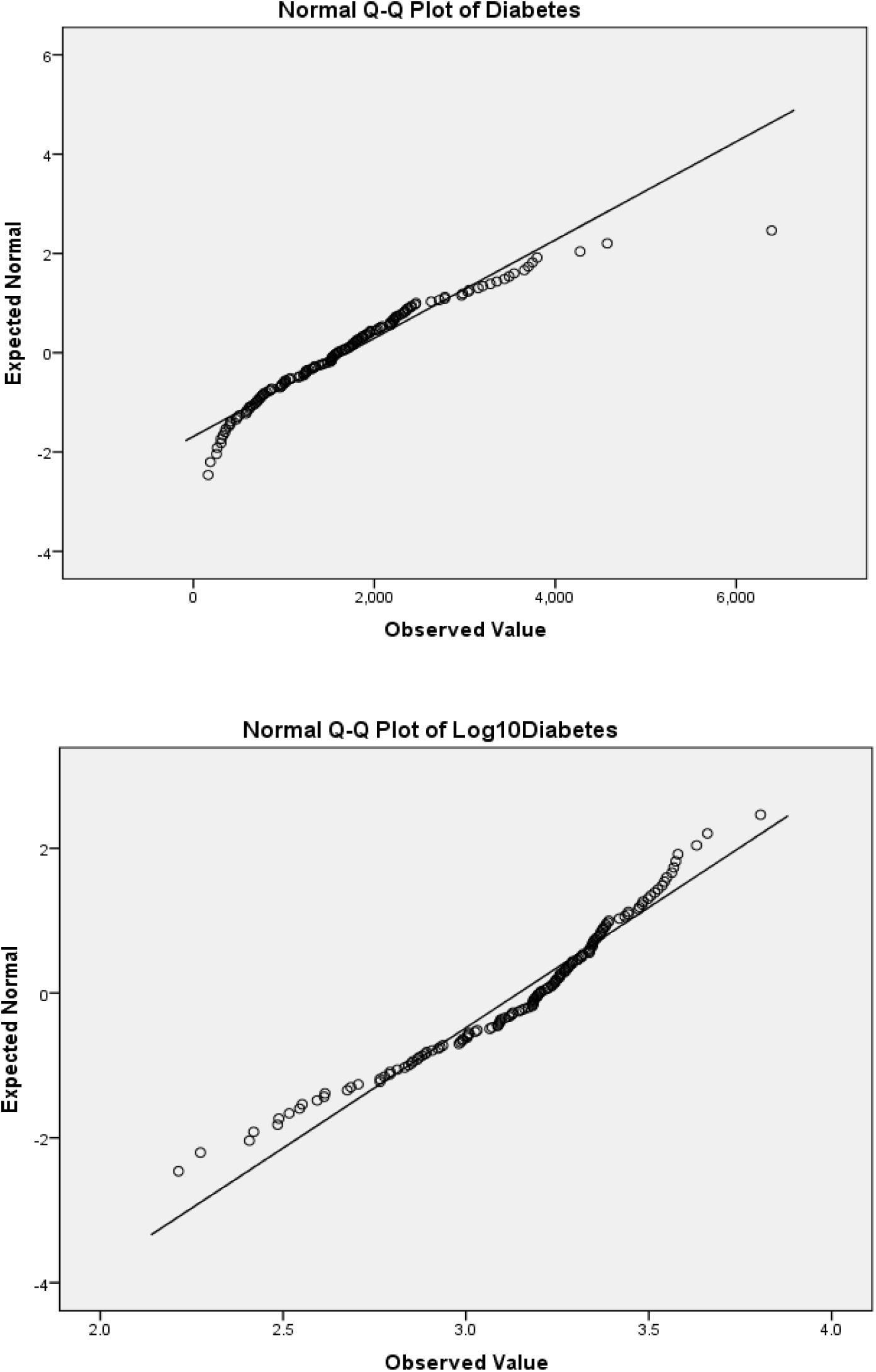
Results of QQ-plots showing non-Gaussian distribution of type 2 diabetes crude rates (top panel) and Gaussian distribution of logged type 2 diabetes crude rates (bottom panel)

**Supplementary Figure 2.**
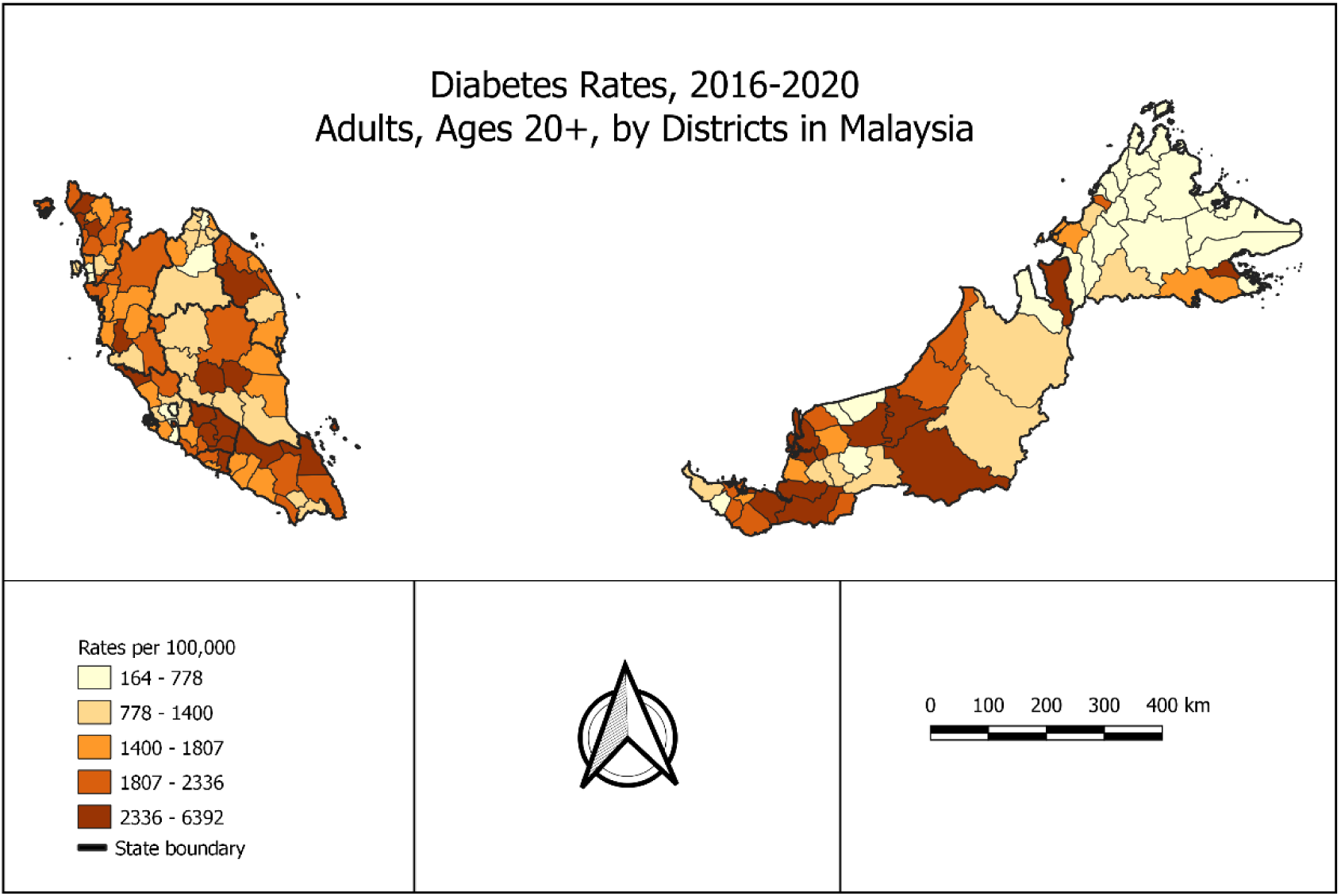
Results of quantile map showing untransformed crude rates of type 2 diabetes per 100,000 population by administrative districts in Malaysia

**Supplementary Figure 3.**
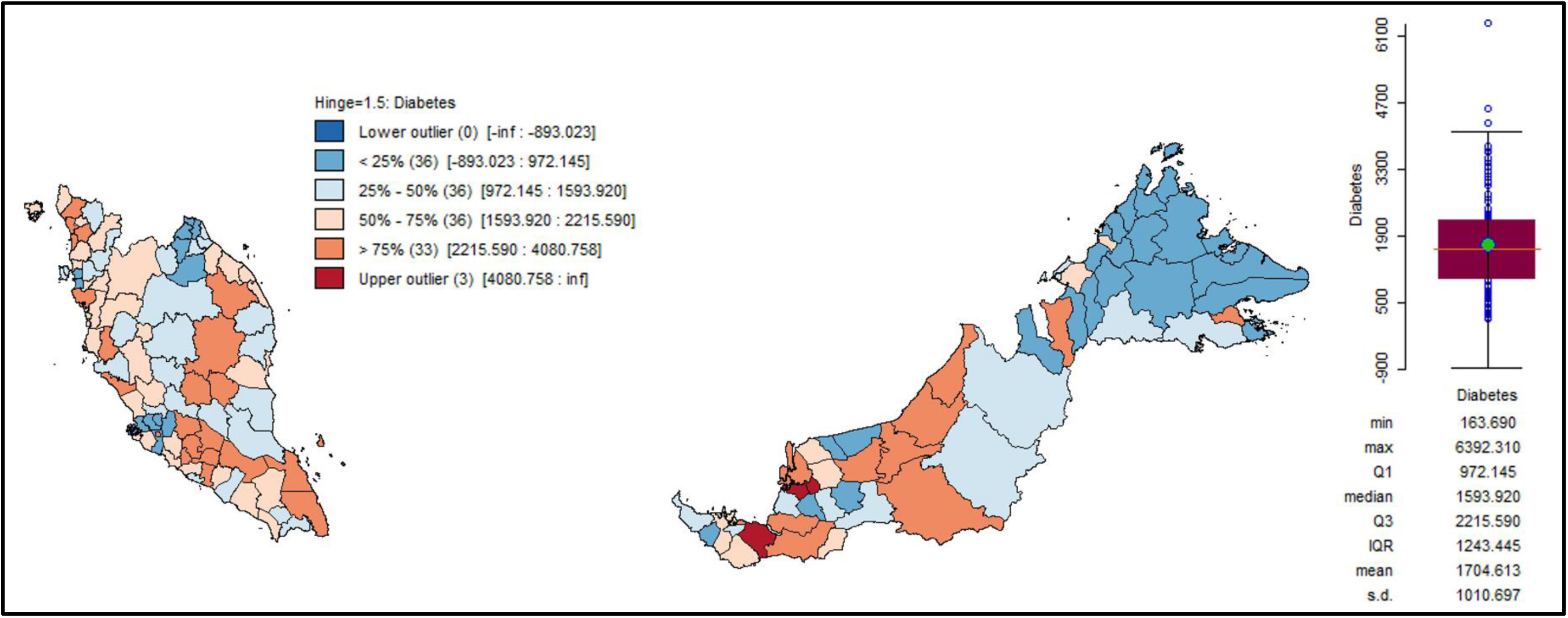
Results showing a box map – boxplot geostatistical summary of the distribution of type 2 diabetes crude rates per 100,000 population among adults by administrative districts in Malaysia.

**Supplementary Figure 4.**
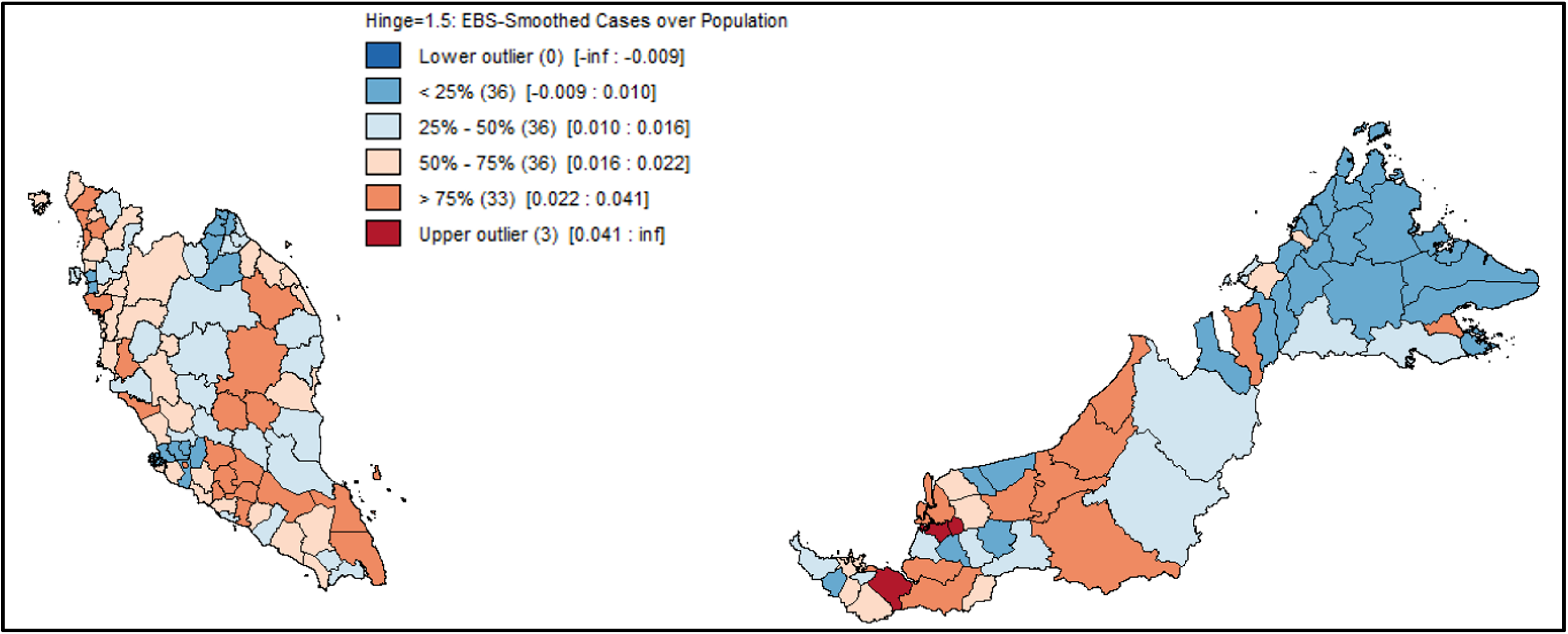
Results showing an Empirical Bayes smoothed rates map of type 2 diabetes distribution by administrative districts in Malaysia.

**Supplementary Figure 5.**
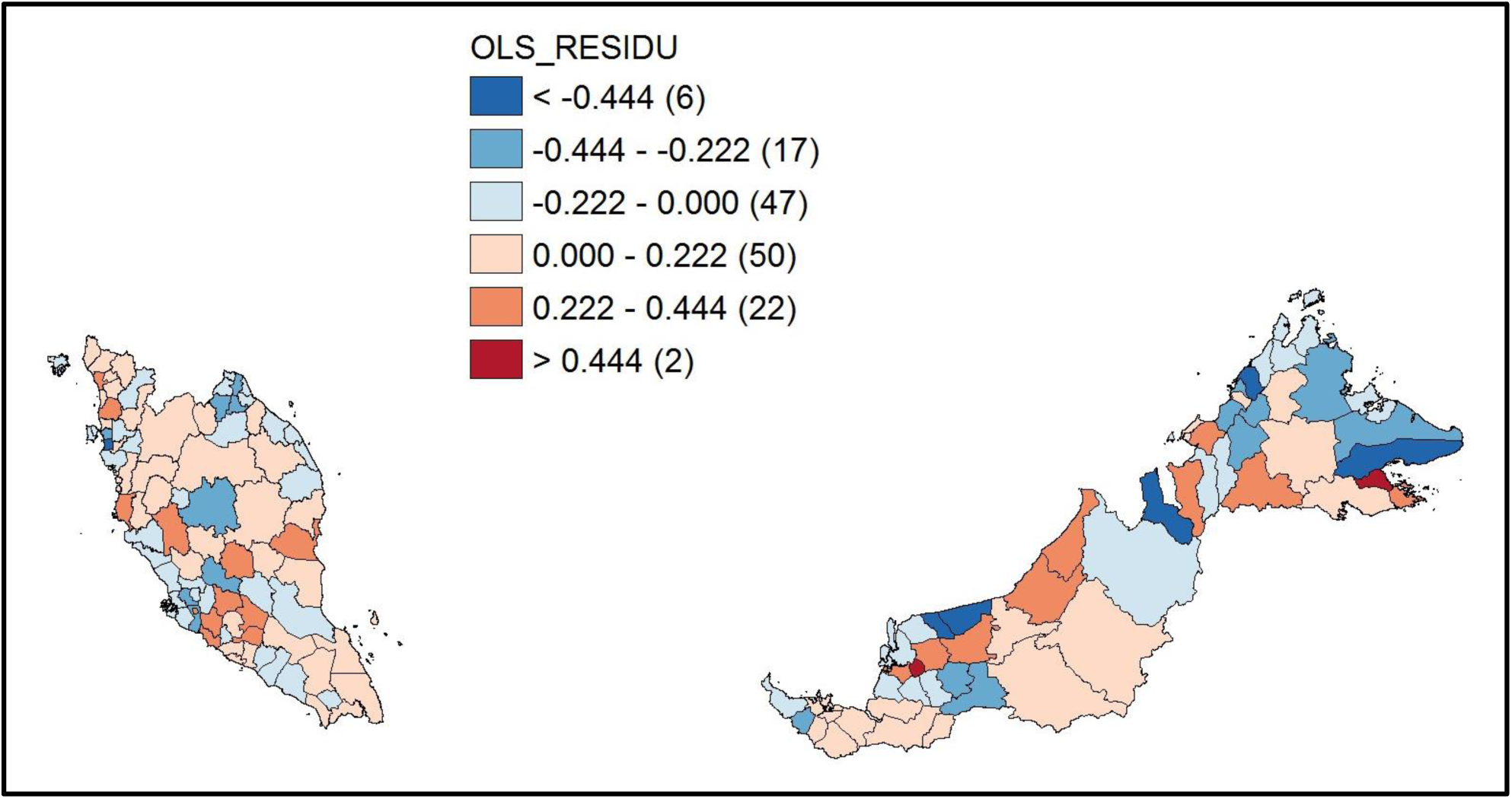
Results showing a standard deviation map of the Ordinary Least Squares Regression (OLS) residuals by administrative districts in Malaysia.

**Supplementary Figure 6.**
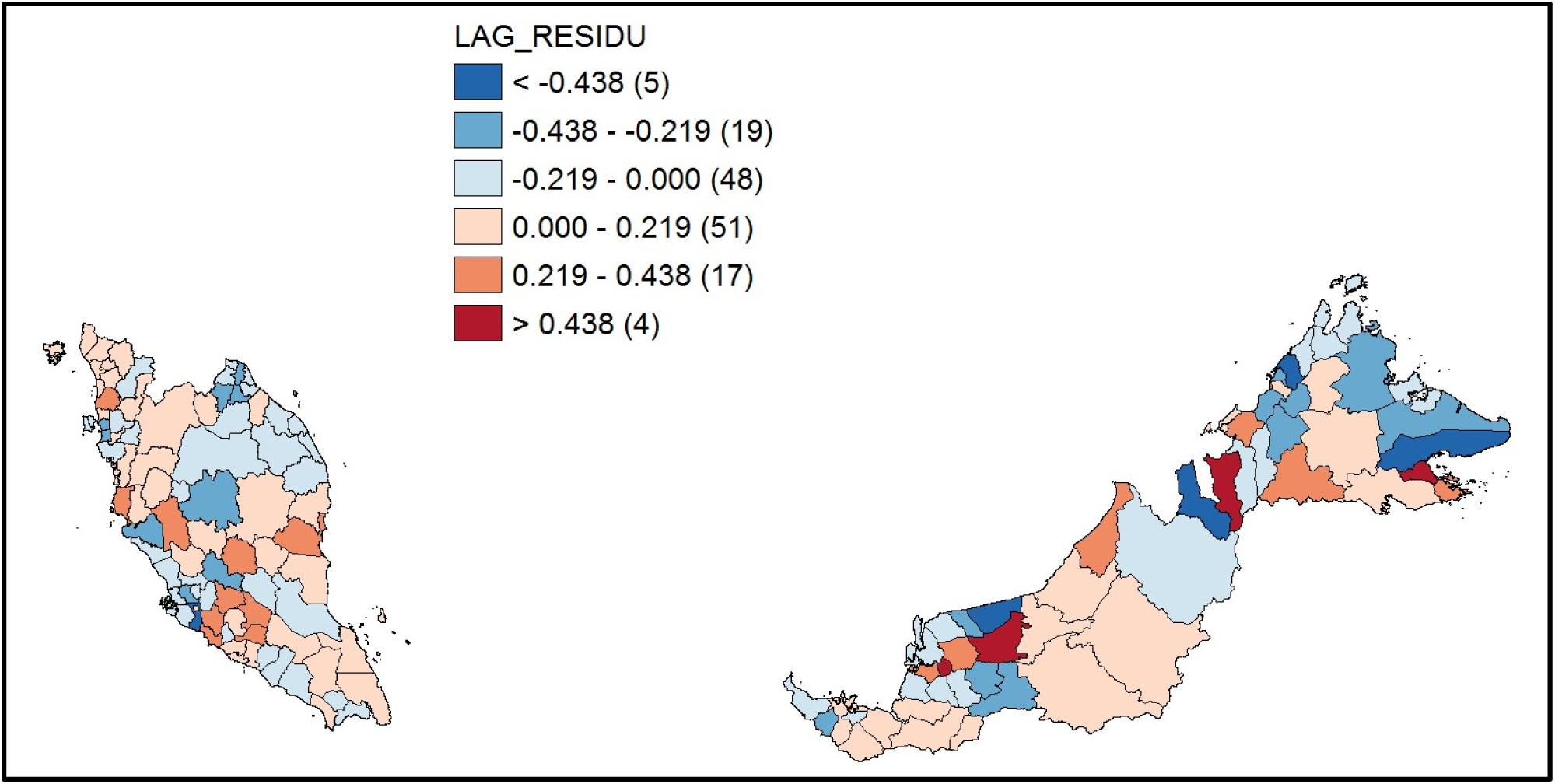
Results showing a standard deviation map of the Spatial Lag Regression (SLM) model residuals by administrative districts in Malaysia.

**Supplementary Figure 7.**
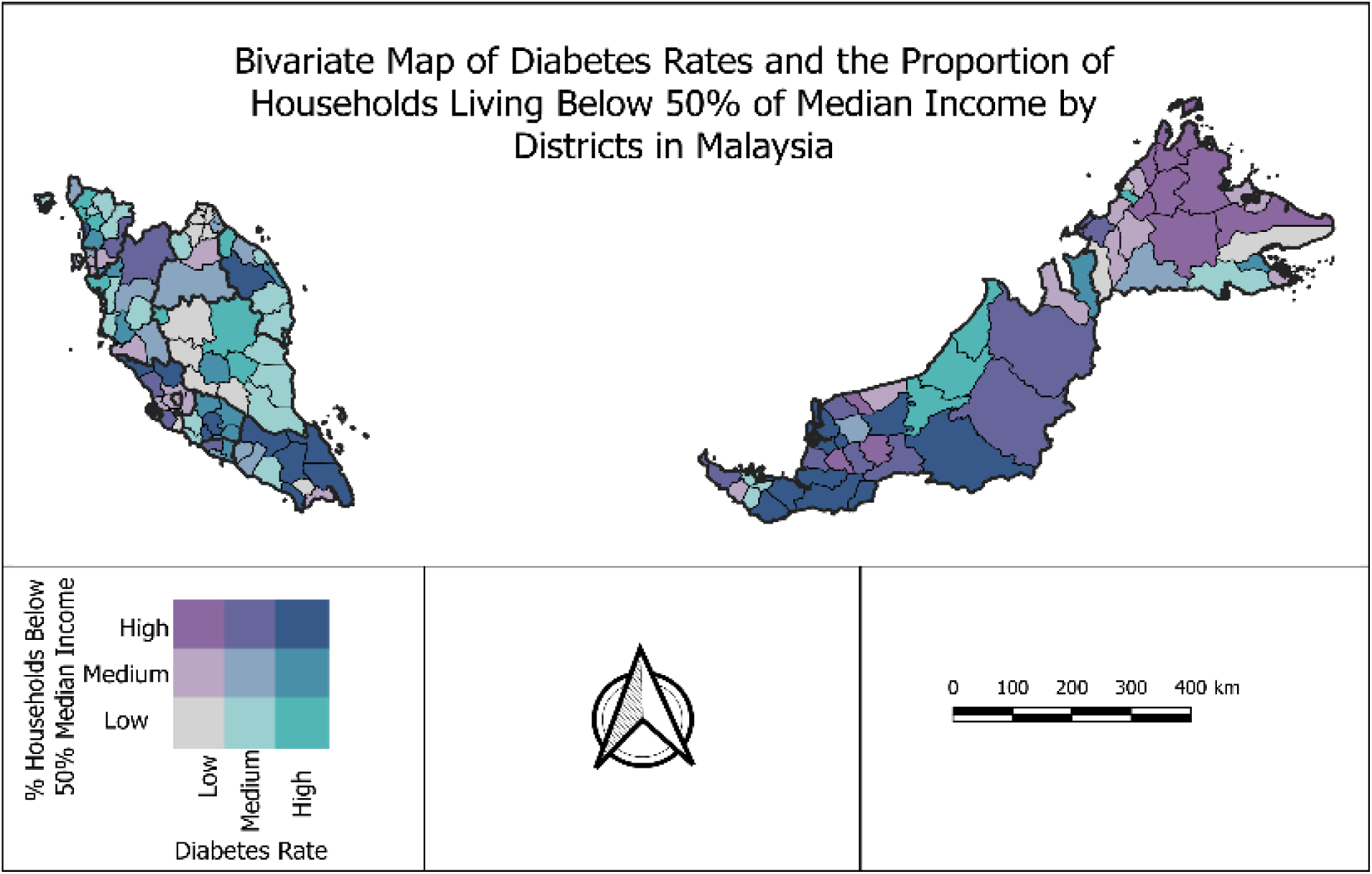
Supplementary figure 7 demarcates a bivariate choropleth map on the associations between diabetes rates and the proportion of households living below 50% of median income by administrative districts in Malaysia. Districts within the states of Johor (Kota Tinggi, Pontian, Kluang, Segamat, Mersing), Negeri Sembilan (Kuala Pilah, Rembau), Selangor (Sabak Bernam, Ulu Selangor), Kedah (Yan), Terengganu (Hulu Terengganu), Sabah (Beaufort, Kuala Penyu), and Sarawak (Kapit, Tatau, Daro, Sarikei, Meradong, Betong, Sri Aman, Lubok Antu, Simunjan, Serian) fell into the high-proportion/high-rates group. One district in Sabah (Penampang), three in North Sarawak (Miri, Bintulu, Tatau), some scattered districts in the Central, Northern and East-Coast regions fell into the low-proportion/high-rates diabetes group.

**Supplementary Figure 8.**
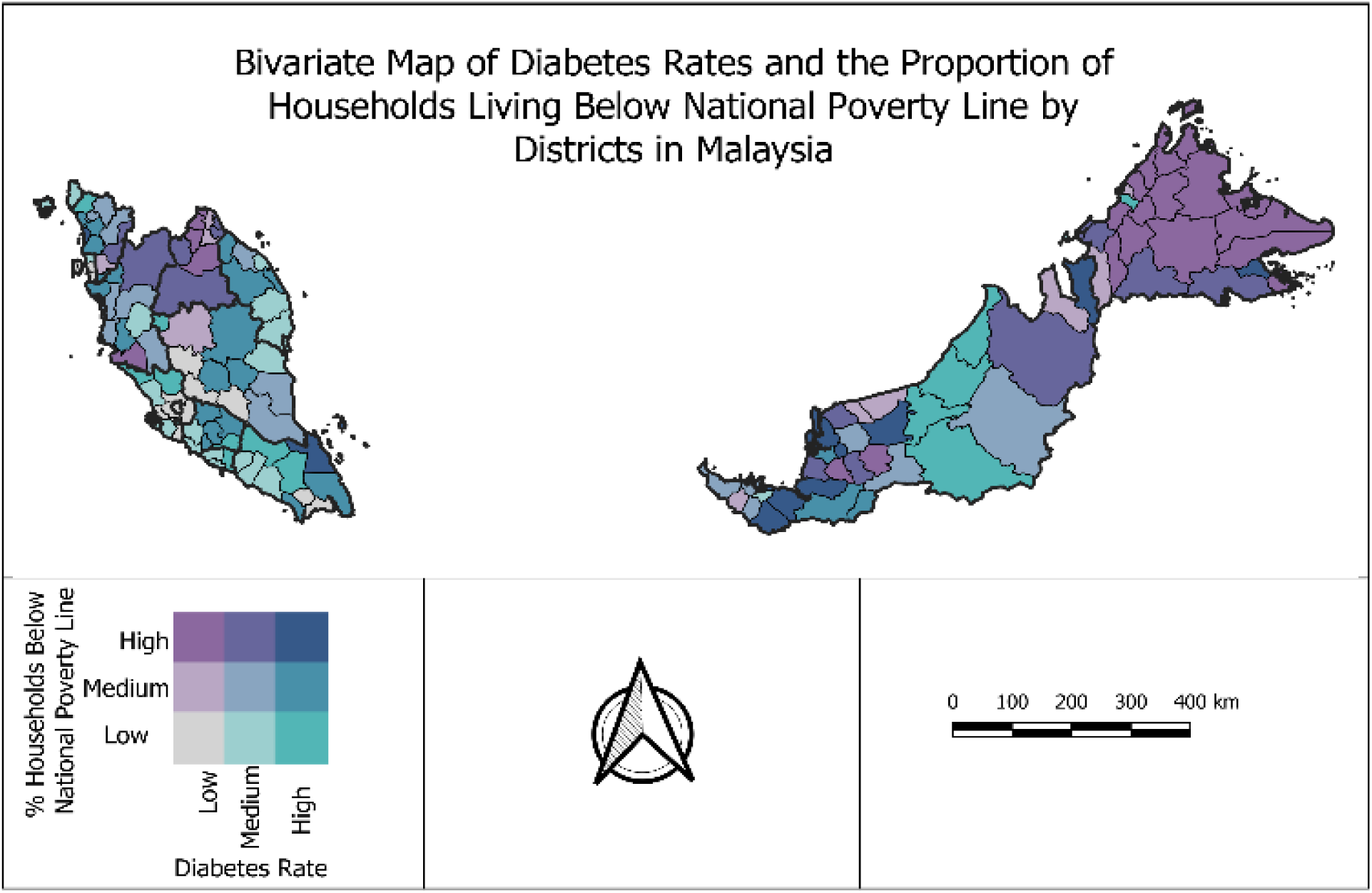
Supplementary figure 8 shows a bivariate choropleth map on the associations between diabetes rates and the proportion of households living below national poverty line by administrative districts in Malaysia. Districts within the states of Johor (Mersing), Kedah (Yan), Sabah (Kunak), and Sarawak (Lawas, Selangau, Daro, Meradong, Betong, Simunjan, Serian) fell into the high-proportion/high-rates group. Relatively sparse districts within the Southern, Central, and Northern regions plus four districts in Sarawak (Miri, Bintulu, Tatau, Kapit) and one in Sabah (Penampang) fell into the low-proportion/high-rates diabetes group.

**Supplementary Figure 9.**
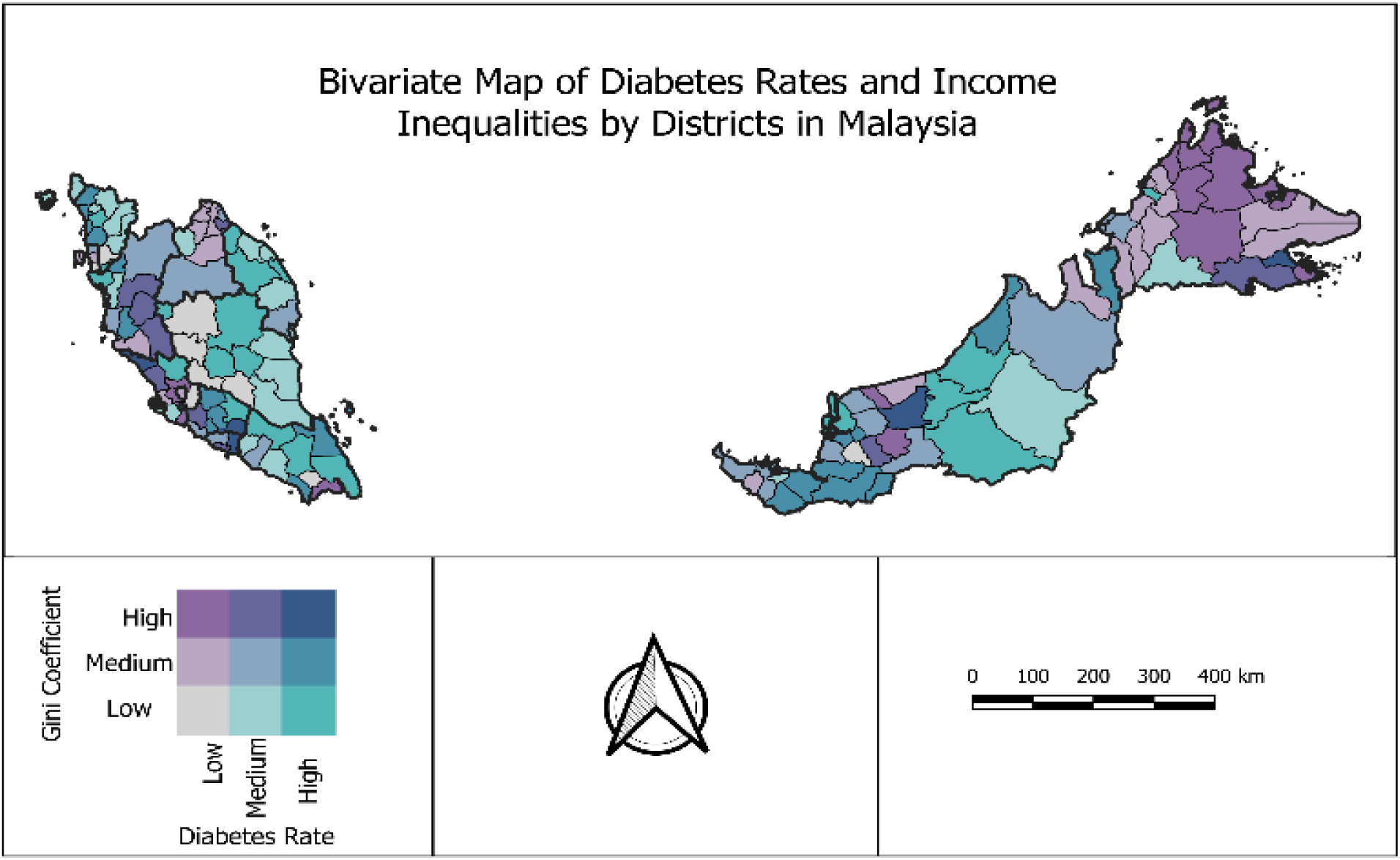
Supplementary figure 9 exhibits a bivariate choropleth map on the associations between diabetes rates and income inequalities (measured by Gini coefficient) by administrative districts in Malaysia. Districts within the states of Negeri Sembilan (Tampin), Melaka (Jasin), Selangor (Sabak Bernam), Sabah (Kunak), and Sarawak (Selangau) fell into the high-inequalities (third tertile)/high-rates group. Most districts within the Southern, Central, East Coast, and Northern regions plus districts in the North, East, and South of Sarawak and one in Sabah (Penampang) fell into the low-inequalities (first tertile)/high-rates diabetes group.

**Supplementary Figure 10.**
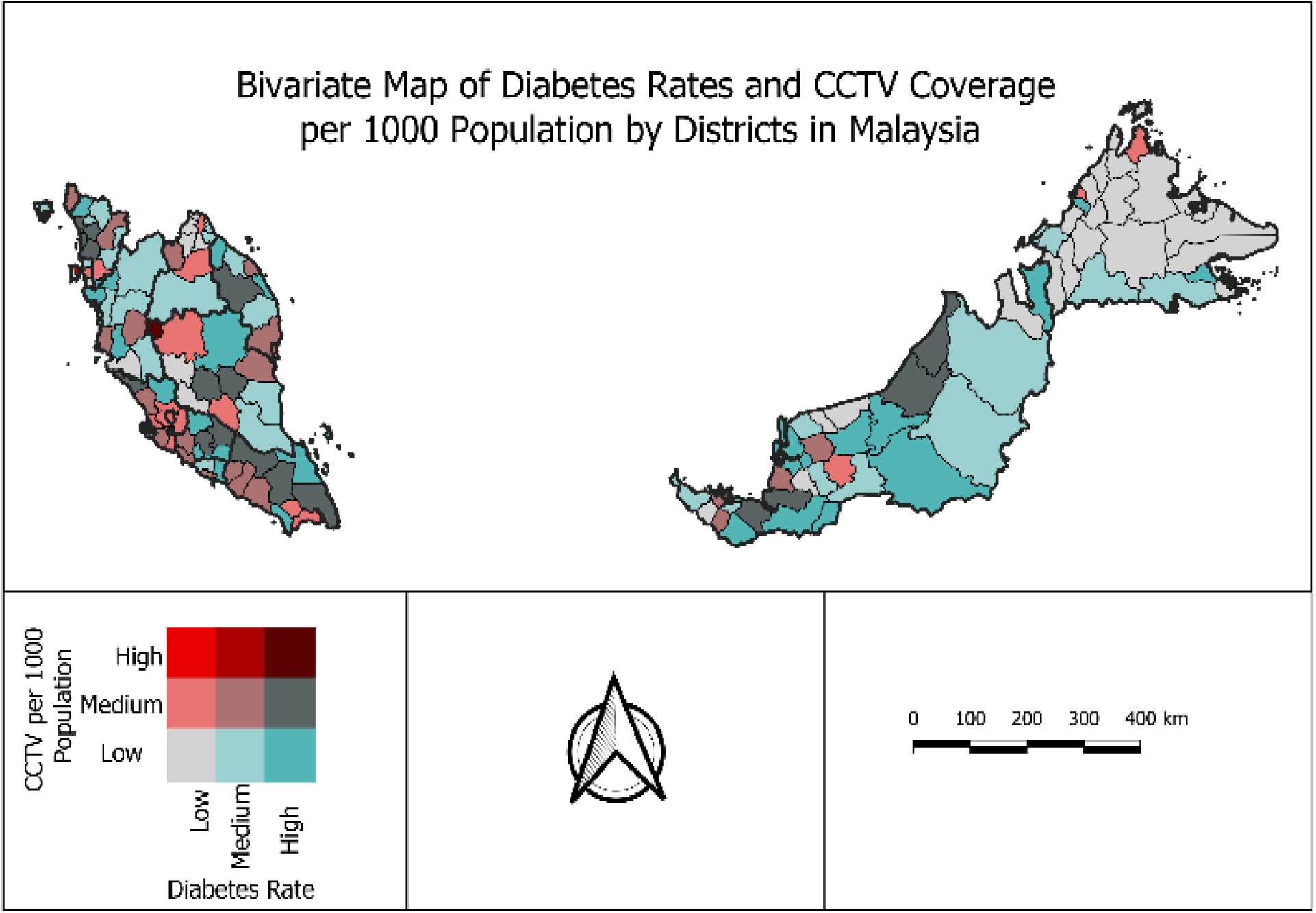
Supplementary figure 10 shows a bivariate choropleth map on the associations between diabetes rates and CCTV coverage per 1000 population by administrative districts in Malaysia. High CCTV coverage/high-diabetes rates were mainly focused in two major points, namely the districts of Cameron Highlands (Pahang) and Timur Laut (Pulau Pinang). Diabetes rates were relatively high across most districts in Peninsular Malaysia and the state of Sarawak, coherently these districts had relatively low CCTV coverage per 1000 population.

**Supplementary Figure 11.**
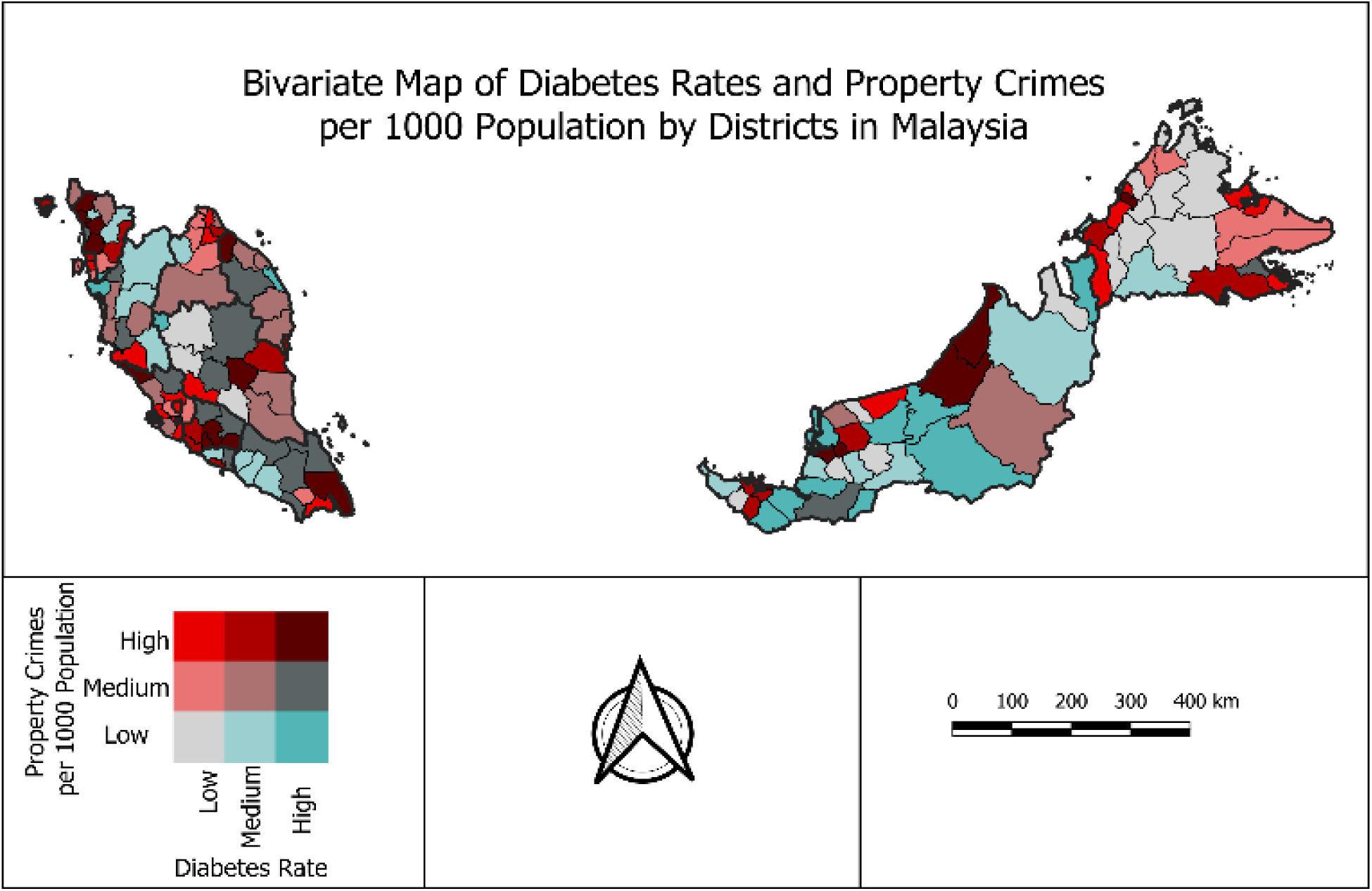
Supplementary figure 11 shows a bivariate choropleth map on the associations between diabetes rates and property crimes per 1000 population by administrative districts in Malaysia. Districts with high property crimes/high-diabetes rates were found in Johor (Kota Tinggi), Negeri Sembilan (Tampin, Kuala Pilah, Rembau), Pahang (Maran), Selangor (Sabak Bernam), Terengganu (Besut), Kedah (Kubang Pasu, Kota Setar, Pendang, Kuala Muda), Sabah (Penampang), and Sarawak (Miri, Bintulu, Sarikei, Meradong). Consistently, diabetes rates were relatively high across districts in Kerian (Perak), Bandar Baharu (Kedah), Cameron Highlands (Pahang), Marang (Terengganu), and Lawas, Kapit, Tatau, Selangau, Daro, Betong, Lubok Antu, Simunjan, Serian (Sarawak) where property crimes were relatively low.

**Supplementary Figure 12.**
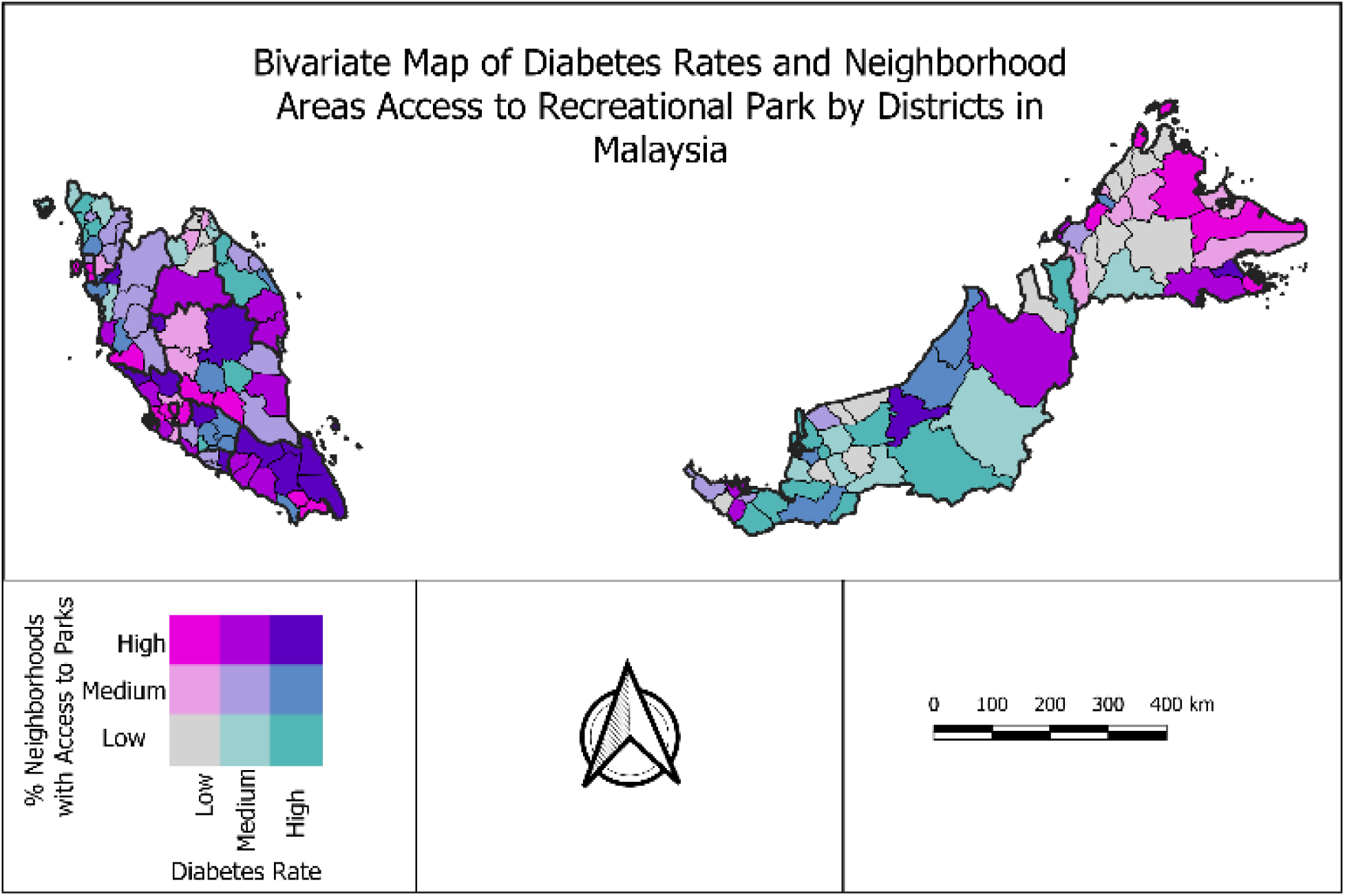
Supplementary figure 12 exhibits a bivariate choropleth map on the associations between diabetes rates and neighbourhood areal access to recreational parks by administrative districts in Malaysia. Districts with high percentage access/high-diabetes rates were found in Johor (Kota Tinggi, Mersing, Kluang, Segamat), Melaka (Jasin), Negeri Sembilan (Jelebu), Pahang (Cameron Highlands, Jerantut), Selangor (Ulu Selangor, Sabak Bernam), Perak (Kampar), Sabah (Kunak), and Sarawak (Tatau). Diabetes rates were relatively high across districts in Rembau (Negeri Sembilan), Maran (Pahang), Besut and Hulu Terengganu (Terengganu), Kubang Pasu, Kota Setar, Yan, and Pendang (Kedah), and Lawas, Kapit, Selangau, Daro, Betong, Lubok Antu, Simunjan, Serian, and Sarikei (Sarawak) where access to recreational parks were low.

**Supplementary Figure 13.**
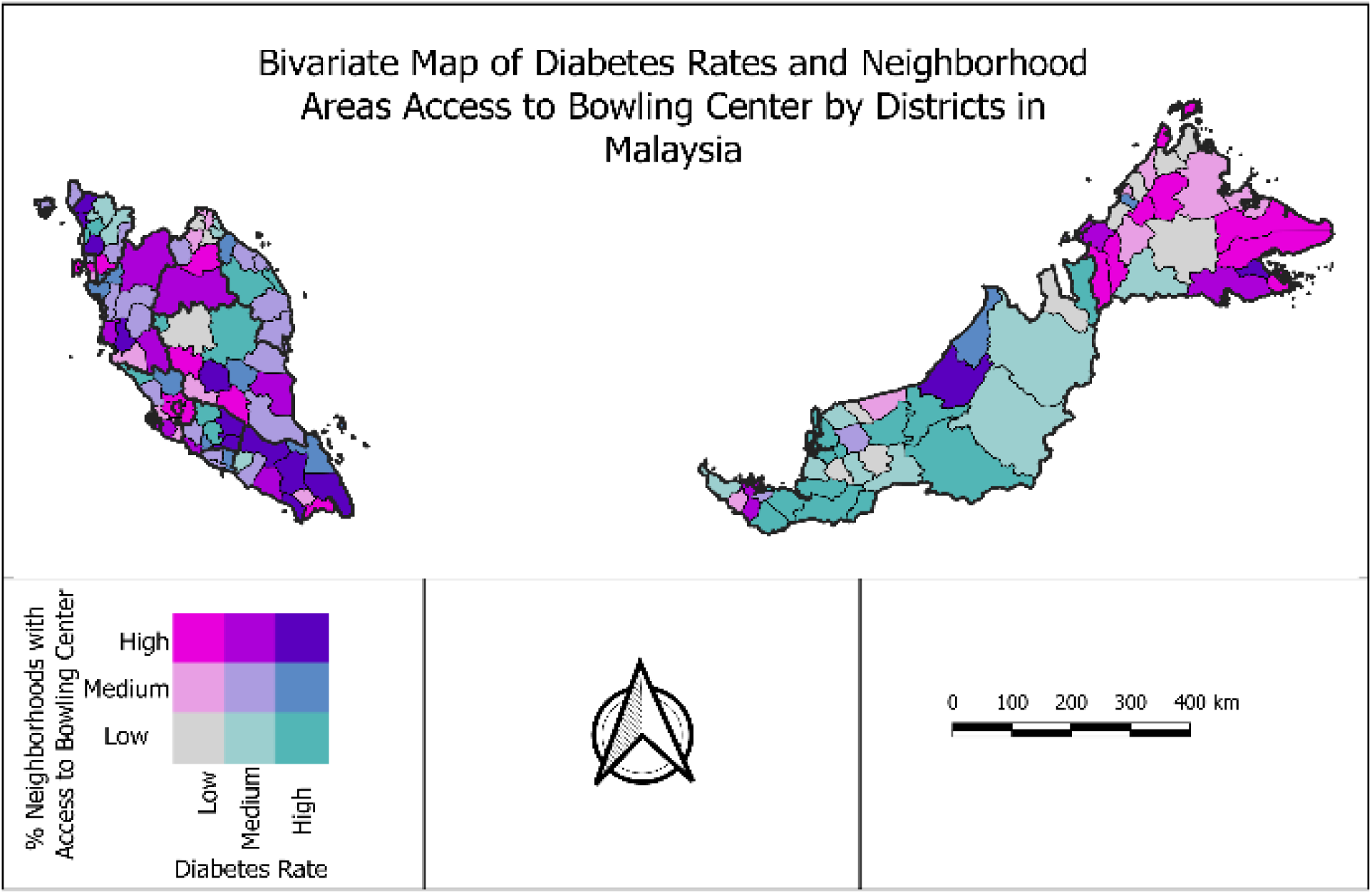
Supplementary figure 13 exhibits a bivariate choropleth map on the associations between diabetes rates and neighbourhood areal access to bowling centers by administrative districts in Malaysia. Districts with high percentage access/high-diabetes rates were found in Johor (Kota Tinggi, Pontian, Kluang, Segamat), Negeri Sembilan (Jempol, Tampin), Pahang (Temerloh), Perak (Batang Padang), Kedah (Kuala Muda, Kota Setar, Kubang Pasu), Sabah (Kunak), and Sarawak (Bintulu). Diabetes rates were relatively high across districts in Jelebu and Kuala Pilah (Negeri Sembilan), Sabak Bernam (Selangor), Cameron Highlands and Jerantut (Pahang), Yan and Pendang (Kedah), Marang and Hulu Terengganu (Terengganu), Lawas, Kapit, Selangau, Daro, Betong, Lubok Antu, Simunjan, Serian, Tatau, Meradong, Sri Aman, and Sarikei (Sarawak) where access to bowling centers were low.

**Supplementary Table 1.** Results of univariate ordinary least squares regression with variation inflation factor (VIF) values for variable selection into the multivariate OLS model.

| Characteristics | $\beta$ | SE | p-Value | VIF |
| --- | --- | --- | --- | --- |
| Proportion of women (%) | 5.541 | 2.510 | 0.029 | 3.417 |
| Proportion of Bumiputera (%) | 0.879 | 0.599 | 0.145 | 4.088 |
| Proportion of Chinese (%) | 0.811 | 0.600 | 0.179 | 17.530 |
| Proportion of Indian (%) | 0.315 | 0.844 | 0.710 | 4.689 |
| Proportion of adults aged 35-49 years (%) | -1.550 | 1.156 | 0.183 | 3.775 |
| Proportion of adults aged 50-64 years (%) | 1.429 | 0.035 | 0.834 | 4.069 |
| Proportion of adults aged $\geq 65$ years (%) | 1.429 | 0.068 | 0.604 | 3.873 |
| Proportion of households living below fifty percent of median income (%) | 0.003 | 0.259 | 0.034 | 3.268 |
| Proportion of households living below national poverty line (%) | 0.004 | -0.406 | 0.003 | 3.916 |
| Income inequalities (Gini coefficient) | 0.796 | -0.195 | 0.042 | 2.006 |
| Unemployment rate (%) | 0.003 | 0.052 | 0.473 | 1.190 |
| Urban growth rate (%) | 0.003 | 0.053 | 0.476 | 1.200 |
| Population density | 0.00006 | 0.000001 | 0.049 | 5.620 |
| CCTV coverage per 1000 population | 0.064 | 0.034 | 0.067 | 2.044 |
| Property crimes per 1000 population | 0.023 | 0.008 | 0.106 | 2.347 |
| Drug addicts per 1000 population | 0.020 | 0.021 | 0.543 | 3.466 |
| Proportion of population not engaged in fitness/ exercise or sports activities (%) | -0.226 | 0.125 | 0.072 | 1.920 |
| % of area with access to parks<sup>#</sup> | 0.007 | 0.003 | 0.011 | 4.246 |
| % of area with access to jogging track<sup>#</sup> | -0.003 | 0.002 | 0.135 | 4.634 |
| % of area with access to cycling track<sup>#</sup> | -0.00017 | 0.001 | 0.990 | 4.108 |
| % of area with access to mini stadium<sup>#</sup> | 0.003 | 0.002 | 0.104 | 2.200 |
| % of area with access to football field<sup>#</sup> | 0.002 | 0.003 | 0.534 | 2.894 |
| % of area with access to gymnasium<sup>#</sup> | 0.001 | 0.0001 | 0.881 | 3.518 |
| % of area with access to bowling centre<sup>#</sup> | -0.003 | 0.001 | 0.046 | 2.484 |

**Note:** here *p*-values (\*\*) denotes strong evidence of statistical significance, (\*) denotes moderate evidence of statistical significance; (#) denotes less than five kilometres of proximity from neighbourhoods; $\beta$ indicates unstandardized coefficients; SE indicates standard error; VIF indicates variation inflation factor; Reference group for gender was the proportion of men, for ethnicity was the proportion of other ethnicities, and for age was the proportion of adults aged 20-34 years.

**Supplementary Table 2.** Results of two-way interaction analyses adjusted for spatial correlation between access to fitness facilities with neighborhood poverty and inequity indicators from spatial lag model (SLM).

| **Models** | **Neighborhood indicators** | **$\beta$** | **SE** | ***p*-value** |
| --- | --- | --- | --- | --- |
| 1 | Below 50% median income | 0.003 | 0.003 | 0.251 |
|  | Income inequalities (Gini coefficient) | -2.550 | 0.743 | 0.001 |
|  | % of area with access to parks | 0.005 | 0.001 | <0.001 |
|  | % of area with access to bowling centre | -0.005 | 0.001 | <0.001 |
| 2 | Below poverty line | -0.009 | 0.003 | <0.001 |
|  | Income inequalities (Gini coefficient) | -0.935 | 0.680 | 0.169 |
|  | % of area with access to parks | 0.003 | 0.001 | 0.054 |
|  | % of area with access to bowling centre | -0.005 | 0.001 | <0.001 |
| 3 | Below 50% median income | 0.004 | 0.003 | 0.134 |
|  | Income inequalities (Gini coefficient) | -6.952 | 2.229 | 0.002 |
|  | % of area with access to parks | -0.018 | 0.011 | 0.106 |
|  | % of area with access to bowling centre | -0.005 | 0.001 | <0.001 |
|  | Interaction (access to parks*Gini coefficient) | 0.063 | 0.030 | 0.037 |
| 4 | Below poverty line | -0.013 | 0.005 | 0.011 |
|  | Income inequalities (Gini coefficient) | -0.936 | 0.678 | 0.168 |
|  | % of area with access to parks | 0.002 | 0.002 | 0.298 |
|  | % of area with access to bowling centre | -0.005 | 0.001 | <0.001 |
|  | Interaction (access to parks*below poverty line) | 0.00007 | 0.00007 | 0.346 |
| 5 | Below poverty line | -0.008 | 0.003 | 0.002 |
|  | Income inequalities (Gini coefficient) | -3.455 | 2.193 | 0.115 |
|  | % of area with access to parks | -0.010 | 0.011 | 0.346 |
|  | % of area with access to bowling centre | -0.005 | 0.001 | <0.001 |
|  | Interaction (access to parks*Gini) | 0.036 | 0.030 | 0.226 |
| 6 | Below 50% median income | 0.005 | 0.003 | 0.103 |
|  | Income inequalities (Gini coefficient) | -2.592 | 0.740 | <0.001 |
|  | % of area with access to parks | 0.005 | 0.001 | <0.001 |
|  | % of area with access to bowling centre | -0.003 | 0.002 | 0.217 |
|  | Interaction (access to bowling*below 50% median income) | -0.0001 | 0.0001 | 0.230 |
| 7 | Below 50% median income | 0.003 | 0.003 | 0.296 |
|  | Income inequalities (Gini coefficient) | -1.790 | 1.003 | 0.074 |
|  | % of area with access to parks | 0.005 | 0.001 | <0.001 |
|  | % of area with access to bowling centre | 0.007 | 0.010 | 0.508 |
|  | Interaction (access to bowling*Gini) | -0.031 | 0.028 | 0.263 |
| 8 | Below poverty line | -0.009 | 0.003 | 0.010 |
|  | Income inequalities (Gini coefficient) | -0.934 | 0.682 | 0.171 |
|  | % of area with access to parks | 0.003 | 0.001 | 0.055 |
|  | % of area with access to bowling centre | -0.005 | 0.002 | 0.003 |
|  | Interaction (access to bowling*below poverty line) | 0.0000009 | 0.00008 | 0.991 |
| 9 | Below poverty line | -0.008 | 0.003 | 0.001 |
|  | Income inequalities (Gini coefficient) | -0.483 | 0.891 | 0.588 |
|  | % of area with access to parks | 0.003 | 0.001 | 0.044 |
|  | % of area with access to bowling centre | 0.003 | 0.010 | 0.759 |
|  | Interaction (access to bowling*Gini) | -0.021 | 0.027 | 0.433 |
| 10 | Below poverty line | -0.010 | 0.002 | <0.001 |
|  | % of area with access to parks | 0.003 | 0.001 | 0.069 |
|  | % of area with access to bowling centre | -0.005 | 0.001 | <0.001 |

**Supplementary Table 3.** Results of two-way interaction analyses adjusted for spatial correlation between access to fitness facilities with neighborhood safety and inequity indicators from spatial lag model (SLM).

| **Models** | **Neighborhood indicators** | **$\beta$** | **SE** | ***p*-value** |
| --- | --- | --- | --- | --- |
| 1 | CCTV per 1000 population | 0.044 | 0.027 | 0.095 |
|  | Income inequalities (Gini coefficient) | -1.972 | 0.611 | <0.001 |
|  | % of area with access to parks | 0.004 | 0.001 | 0.003 |
|  | % of area with access to bowling centre | -0.005 | 0.001 | <0.001 |
| 2 | Property crimes per 1000 population | 0.014 | 0.007 | 0.061 |
|  | Income inequalities (Gini coefficient) | -2.141 | 0.609 | <0.001 |
|  | % of area with access to parks | 0.004 | 0.001 | 0.002 |
|  | % of area with access to bowling centre | -0.005 | 0.001 | <0.001 |
| 3 | CCTV per 1000 population | 0.224 | 0.256 | 0.380 |
|  | Income inequalities (Gini coefficient) | -1.939 | 0.610 | 0.001 |
|  | % of area with access to parks | 0.004 | 0.001 | 0.003 |
|  | % of area with access to bowling centre | -0.005 | 0.001 | <0.001 |
|  | Interaction (access to parks*CCTV) | -0.002 | 0.003 | 0.480 |
| 4 | CCTV per 1000 population | 0.048 | 0.026 | 0.069 |
|  | Income inequalities (Gini coefficient) | -5.943 | 2.092 | 0.005 |
|  | % of area with access to parks | -0.017 | 0.011 | 0.113 |
|  | % of area with access to bowling centre | -0.005 | 0.001 | <0.001 |
|  | Interaction (access to parks*Gini) | 0.059 | 0.030 | 0.047 |
| 5 | Property crimes per 1000 population | 0.038 | 0.022 | 0.084 |
|  | Income inequalities (Gini coefficient) | -2.-71 | 0.610 | <0.001 |
|  | % of area with access to parks | 0.006 | 0.002 | 0.001 |
|  | % of area with access to bowling centre | -0.005 | 0.001 | <0.001 |
|  | Interaction (access to parks*property crimes) | -0.0006 | 0.0005 | 0.243 |
| 6 | Property crimes per 1000 population | 0.014 | 0.007 | 0.062 |
|  | Income inequalities (Gini coefficient) | -5.828 | 2.088 | 0.005 |
|  | % of area with access to parks | -0.015 | 0.011 | 0.154 |
|  | % of area with access to bowling centre | -0.005 | 0.001 | <0.001 |
|  | Interaction (access to parks*Gini) | 0.055 | 0.030 | 0.065 |
| 7 | CCTV per 1000 population | -0.008 | 0.054 | 0.884 |
|  | Income inequalities (Gini coefficient) | -2.073 | 0.615 | <0.001 |
|  | % of area with access to parks | 0.004 | 0.001 | 0.003 |
|  | % of area with access to bowling centre | -0.005 | 0.001 | <0.001 |
|  | Interaction (access to bowling*CCTV) | 0.001 | 0.001 | 0.269 |
| 8 | CCTV per 1000 population | 0.046 | 0.026 | 0.081 |
|  | Income inequalities (Gini coefficient) | -1.137 | 0.878 | 0.196 |
|  | % of area with access to parks | 0.004 | 0.001 | 0.002 |
|  | % of area with access to bowling centre | 0.008 | 0.010 | 0.409 |
|  | Interaction (access to bowling*Gini) | -0.036 | 0.027 | 0.190 |
| 9 | Property crimes per 1000 population | 0.015 | 0.008 | 0.063 |
|  | Income inequalities (Gini coefficient) | -2.134 | 0.610 | <0.001 |
|  | % of area with access to parks | 0.004 | 0.001 | 0.001 |
|  | % of area with access to bowling centre | -0.005 | 0.001 | 0.001 |
|  | Interaction (access to bowling*property crimes) | -0.0001 | 0.0005 | 0.765 |
| 10 | Property crimes per 1000 population | 0.013 | 0.007 | 0.069 |
|  | Income inequalities (Gini coefficient) | -1.420 | 0.878 | 0.106 |
|  | % of area with access to parks | 0.004 | 0.001 | 0.001 |
|  | % of area with access to bowling centre | 0.007 | 0.010 | 0.518 |
|  | Interaction (access to bowling*Gini) | -0.031 | 0.027 | 0.257 |

**Supplementary Table 4.** Results of two-way interaction analyses adjusted for spatial correlation between access to fitness facilities with neighborhood poverty and safety indicators from spatial lag model (SLM).

| **Models** | **Neighborhood indicators** | **$\beta$** | **SE** | ***p*-value** |
| --- | --- | --- | --- | --- |
| 1 | CCTV per 1000 population | 0.031 | 0.026 | 0.229 |
|  | Below poverty line | -0.010 | 0.002 | <0.001 |
|  | % of area with access to parks | 0.002 | 0.001 | 0.090 |
|  | % of area with access to bowling centre | -0.005 | 0.001 | <0.001 |
| 2 | Property crimes per 1000 population | 0.008 | 0.007 | 0.271 |
|  | Below poverty line | -0.010 | 0.002 | <0.001 |
|  | % of area with access to parks | 0.003 | 0.001 | 0.067 |
|  | % of area with access to bowling centre | -0.005 | 0.001 | <0.001 |
| 3 | CCTV per 1000 population | 0.155 | 0.253 | 0.540 |
|  | Below poverty line | -0.010 | 0.002 | <0.001 |
|  | % of area with access to parks | 0.003 | 0.001 | 0.080 |
|  | % of area with access to bowling centre | -0.005 | 0.001 | <0.001 |
|  | Interaction (access to parks*CCTV) | -0.001 | 0.003 | 0.624 |
| 4 | CCTV per 1000 population | 0.037 | 0.026 | 0.164 |
|  | Below poverty line | -0.015 | 0.005 | 0.002 |
|  | % of area with access to parks | 0.001 | 0.002 | 0.489 |
|  | % of area with access to bowling centre | -0.005 | 0.001 | <0.001 |
|  | Interaction (access to parks*below poverty line) | 0.00009 | 0.00007 | 0.246 |
| 5 | Property crimes per 1000 population | 0.060 | 0.021 | 0.005 |
|  | Below poverty line | -0.011 | 0.002 | <0.001 |
|  | % of area with access to parks | 0.005 | 0.002 | 0.003 |
|  | % of area with access to bowling centre | -0.005 | 0.001 | <0.001 |
|  | Interaction (access to parks*property crimes) | -0.001 | 0.0005 | 0.010 |
| 6 | Property crimes per 1000 population | 0.008 | 0.007 | 0.263 |
|  | Below poverty line | -0.014 | 0.005 | 0.004 |
|  | % of area with access to parks | 0.002 | 0.002 | 0.341 |
|  | % of area with access to bowling centre | -0.005 | 0.001 | <0.001 |
|  | Interaction (access to parks*below poverty line) | 0.00007 | 0.00007 | 0.340 |
| 7 | CCTV per 1000 population | 0.006 | 0.053 | 0.907 |
|  | Below poverty line | -0.010 | 0.002 | <0.001 |
|  | % of area with access to parks | 0.002 | 0.001 | 0.087 |
|  | % of area with access to bowling centre | -0.005 | 0.001 | <0.001 |
|  | Interaction (access to bowling*CCTV) | 0.001 | 0.001 | 0.580 |
| 8 | CCTV per 1000 population | 0.032 | 0.026 | 0.223 |
|  | Below poverty line | -0.010 | 0.003 | 0.007 |
|  | % of area with access to parks | 0.002 | 0.001 | 0.096 |
|  | % of area with access to bowling centre | -0.005 | 0.002 | 0.001 |
|  | Interaction (access to bowling*below poverty line) | 0.00002 | 0.00008 | 0.817 |
| 9 | Property crimes per 1000 population | 0.012 | 0.008 | 0.104 |
|  | Below poverty line | -0.011 | 0.002 | <0.001 |
|  | % of area with access to parks | 0.003 | 0.001 | 0.057 |
|  | % of area with access to bowling centre | -0.003 | 0.001 | 0.026 |
|  | Interaction (access to bowling*property crimes) | -0.0008 | 0.0005 | 0.087 |
| 10 | Property crimes per 1000 population | 0.008 | 0.007 | 0.266 |
|  | Below poverty line | -0.010 | 0.003 | <0.001 |
|  | % of area with access to parks | 0.003 | 0.001 | 0.071 |
|  | % of area with access to bowling centre | -0.005 | 0.002 | 0.001 |
|  | Interaction (access to bowling*below poverty line) | -0.00001 | 0.00008 | 0.847 |
| 11 | CCTV per 1000 population | 0.024 | 0.028 | 0.389 |
|  | Below poverty line | -0.010 | 0.002 | <0.001 |
|  | % of area with access to parks | 0.003 | 0.001 | 0.078 |
|  | % of area with access to bowling centre | -0.005 | 0.001 | <0.001 |
|  | Interaction (CCTV*below poverty line) | 0.008 | 0.013 | 0.547 |

